# Investigating the effect of national government physical distancing measures on depression and anxiety during the COVID-19 pandemic through meta-analysis and meta-regression

**DOI:** 10.1101/2020.08.28.20184119

**Authors:** João M. Castaldelli-Maia, Megan E. Marziali, Ziyin Lu, Silvia S. Martins

## Abstract

**Background:** COVID-19 physical distancing measures can potentially increase the likelihood of mental disorders. It is unknown whether these measures are associated with depression and anxiety.

**Objectives:** To investigate meta-analytic global levels of depression and anxiety during the COVID-19 pandemic and how implementation of mitigation strategies (i.e. public transportation closures, stay-at-home orders, etc.) impacted such disorders.

**Data sources:** Pubmed, MEDLINE, Web of Science, BIOSIS Citation Index, Current Content Connect, PsycINFO, CINAHL, medRxiv, and PsyArXiv databases for depression and anxiety prevalences; Oxford Covid-19 Government Response Tracker for the containment and closure policies indexes; Global Burden of Disease Study for previous levels of depression and anxiety.

**Study eligibility criteria:** Original studies conducted during COVID-19 pandemic, which assessed categorical depression and anxiety, using PHQ-9 and GAD-7 scales (cutoff ≥ 10).

**Participants and interventions:** General population, healthcare providers, students, and patients. National physical distancing measures.

**Study appraisal and synthesis methods:** Meta-analysis and meta-regresssion.

**Results:** In total, 226,638 individuals were assessed within the 60 included studies. Global prevalence of both depression and anxiety during COVID-19 pandemic were 24.0% and 21.3%, respectively. There was a wide variance in the prevalence of both anxiety and depression reported in different regions of the world and countries. Asia, and China particularly, had the lowest prevalence of both disorders. Regarding the impact of mitigation strategies on mental health, only public transportation closures increased anxiety prevalence.

**Limitations:** Country-level data on physical distancing measures and previous anxiety/depression may not necessarily reflect local (i.e., city-specific) contexts.

**Conclusions and implications of key findings:** Mental health concerns should not be viewed only as a delayed consequence of the COVID-19 pandemic, but also as a concurrent epidemic. Our data provides support for policy-makers to consider real-time enhanced mental health services, and increase initiatives to foster positive mental health outcomes.

**Systematic review registration number:** https://doi.org/10.17605/OSF.IO/JQGSF

## 1. Introduction

COVID-19 is an unprecedented health emergency, affecting millions of individuals across the globe.^1^ SARS-Coronavirus-2, the virus which causes COVID-19, is transmitted person-to-person via respiratory droplets.^2^ In order to prevent and lessen spread, countries began implementing mitigation strategies, such as: stay-at-home or shelter-in-place orders, international travel constraints, closure of schools and workplaces, and movement limitations^3^. Despite being necessary public health measures, researchers have speculated that these measures could increase feelings of social isolation and loneliness^4^; this is of importance, as previous studies have demonstrated that social isolation could impact the likelihood of mental disorders^5^ and physical health outcomes^6^. As of yet, it still remains unclear to what extent the COVID-19 mitigation strategies could impact mental health. Thus, it is imperative to investigate the levels of mental health disorders and the possible impacts of social distancing measures on mental health outcomes^7^.

Prior to the pandemic, depression and anxiety were the most prevalent mental health disorders in the world^8^. These mental health disorders have also been connected to social isolation during COVID-19 in local studies^9^. During the COVID-19 pandemic, the levels of such disorders have increased. Pappa et al.^10^ conducted a meta-analysis with thirteen studies that included 33,062 healthcare workers during COVID-19, and reported a prevalence of 23.2% and 22.8% of anxiety and depression, respectively. These prevalences are greater than those found in the pre-COVID-19 era.^8^ Several studies have assessed depression and anxiety using scales involving self-reporting during the pandemic.^11-70^ These studies report a wide range of prevalence estimates, which appear to be dependent on the sub-population of interest (i.e., general population, healthcare providers, students, patients), and the geographic location within which the study is focused.^11-70^ There is a need for meta-analytic investigations generating global prevalence measures for both depression and anxiety during the pandemic, with additional exploration via subgroup analysis.

Further, there are mixed findings regarding the effect of mitigation strategies on depression and anxiety during this pandemic. Previous research has demonstrated marked increases in online search trends for mental health topics (i.e., sleep disturbances, negative thoughts, anxiety, suicidal ideation) prior to the implementation of stay-at-home orders in the U.S..^71^ Further, an online qualitative study evaluated focus groups during the beginning of the social distancing measures in the U.K., where they found negative impacts on well-being and mental health after implementation of mitigation strategies.^72^ Individuals who had lower pay, or vulnerable employment, were the most affected.^72^ Thus, the effects of these physical distancing strategies may be time-sensitive. Moreover, there are varying ongoing physical distancing measures (i.e., school closures, workplace closures, public events cancellations, restrictions on the size of gatherings, public transport closures, stay-at-home orders, restrictions on internal movement between cities and regions within a country, and international travel controls) during different periods, depending on the location.^3^ There is a need to explore whether these strategies have lasting impacts on depression and anxiety, taking different time of exposure thresholds to such physical distancing measures into account.

The present study aims to (1) investigate meta-analytic global levels of depression and anxiety during the COVID-19 pandemic, and (2) explore the effects of these mitigation strategies on depression and anxiety.

## 2. Methods

### 2.1 Study design

We first conducted a meta-analysis of studies related to the COVID-19 pandemic which assessed depression and anxiety using PHQ-9 and GAD-7 scales. Subgroup analysis for region of the world, country, type of population, and coverage were also carried out. Then, we collected national data regarding the implementation of physical distancing measures and mitigation strategies,^3^ along with previous levels of anxiety and depression from a global database.^8^ These data were included in meta-regression models for the investigation of time-sensitive effects of mitigation strategies on depression and anxiety, adjusted for previous levels of such disorders and other possible confounders.

### 2.2. Review Guidelines and Registration

This study followed the PRISMA statement for transparent report of systematic reviews and meta-analysis^73^ and MOOSE guidelines for Meta-analysis Of Observational Studies in Epidemiology.^74^ Figure S1 and S2 respectively present PRISMA and MOOSE checklists reporting the page of the manuscript in which we consider that each item was addressed. This study was registered at the Center for Open Science/Open Science Framework.^75^

### 2.2. Search Strategy

We searched Pubmed, MEDLINE, Web of Science, BIOSIS Citation Index, Current Content Connect, PsycINFO, and CINAHL databases. All searches were conducted with an end date of July 29^th^, 2020. Search terms used were: ((sars-cov-2 OR coronavir* OR alphacoronavirus OR betacoronavirus OR COVID OR COVID-19) AND (PHQ-9 or GAD-7)). As this topic is developing quickly, we accessed pre-print servers medRxiv and PsyArXiv using the above search terms. We also searched the WHO database which includes COVID literature (cite) for studies published by the same date, using the following search terms: (PHQ-9 or GAD-7). In addition to MEDLINE, this database also includes WHO COVID, Elsevier, Lanzhou University/CNKI, LILACS, and WPRIM databases.

### 2.3. Screening and Eligibility

We first removed duplicates from our search results. Screening and eligibility were performed by three researchers independently (JMCM, MEM, ZL). Studies that were written in Chinese were screened by two researchers (JMCM, ZL). Disagreement on the inclusion of a study based on the title or abstract resulted in the study being retained for the next screening stage. Reasons for exclusion of full texts were collected and presented in the PRISMA Flow Diagram (Figure 1).

We included studies that reported categorical assessment of anxiety and depression using GAD-7 and PHQ-9 scales during the COVID-19 pandemic. Randomized controlled trials, cohort studies, case-control studies, and cross-sectional studies were included. Pre-prints and letters were included if they described original research.

### 2.4. Data extraction

Data were extracted by two of three independent reviewers (JMCM, MEM, ZL). Descriptive variables extracted were setting (i.e., country), population type (e.g., pregnant women and children), study design (e.g., cohort and case-control), follow-up time, nature of the control group, number of cases, number of controls, age, and gender. Randomized controlled trials, for this review, were treated as cohort studies. The timepoint for data extraction in prospective studies was either before the intervention (i.e., clinical trials) or during the COVID-19 pandemic (i.e., cohort studies). Data was stored in Excel version 16.16.11.

### 2.5. Quality Assessment

The purpose of this appraisal was to assess the methodological quality of the included studies and to determine the extent to which a study has addressed the possibility of bias in its design, conduct and analysis. All studies included in the present systematic review were subjected to the Joanna Briggs Institute Checklist for Analytical Cross-Sectional Studies,^76^ which assesses sample frame, process and size, setting description, data analysis coverage, valid and reliable assessment methods, appropriate statistical analysis, and an adequate response rate.

### 2.6. Measures

Apart from outcome (depression and anxiety) and exposure (physical distancing measures) variables that are further explained, the present study sought the following data from each included study: the number of individuals enrolled in the study; mean age, standard deviation, and minimum/maximum age range of participants (or median and interquartile range); the proportion of women included; whether the study was nationally representative; whether the study was peer-reviewed; format of data collection (i.e. online); and geographic location, including city, state, and country. Subsequently, we collected data on the previous prevalence of depression and anxiety of each country included in this review.^8^

#### 2.6.1. Anxiety and Depression

The Patient Health Questionnaire-9 (PHQ-9)^77^ is a screening instrument for depressive disorders. It is composed of nine basic items based in the DSM-IV diagnostic criteria for major depressive disorder. The questions assess the frequency of depressive symptoms in the last two weeks. The respondents answer on a scale from 0 (not at all) to 3 (nearly every day). Several studies have used the cut-off ≥10 to define clinically relevant depression.^12-17,19,21,24,26,27,29-36,38,40-45,47,50-54,56,57,59,60,62-70^ The Generalized Anxiety Disorder-7 (GAD-7) is a screening instrument for anxiety symptoms.^78^ The GAD-7 is a validated scale which measures anxiety with seven self-rating items on a four-point scale, similarly to PHQ-9. A cut-off ≥ 10 has been used by several studies to define clinically relevant anxiety.^11,14,16,18-32,35-39,41, 42, 44-59, 61-64, 66-69^ Both the PHQ-9 and GAD-7 have excellent psychometric properties.^77,78^

#### 2.6.2. Exposure: Implementation of Physical Distancing Strategies

We collected national data from the Oxford Covid-19 Government Response Tracker.^3^ All containment and closure policies were included in the present study, as follows:

- School closures (0 - no measures; 1 - recommend closing; 2 - require closing only some levels or categories; 3 - require closing all levels);
- Workplace closures (0 - no measures; 1 - recommend closing or recommend work from home; 2 - require closing or work from home for some sectors or categories of workers; 3 - require closing or work from home for all-but essential workplaces);
- Cancellation of public events (0 - no measures; 1 - recommend cancelling; 2 - require cancelling);
- Restrictions on gatherings (0 - no restrictions; 1 - restrictions on very large gatherings above 1000 people; 2 - restrictions on gatherings between 101-1000 people; 3 - restrictions on gatherings between 11-100 people; 4 - restrictions on gatherings of 10 people or less);
- Public transportation closures (0 - no measures; 1 - recommend closing or significantly reduce volume/route/means of transport available; 2 - require closing or prohibit most citizens from using it);
- Stay at home requirements (0 - no measures; 1 - recommend not leaving house; 2 - require not leaving house with exceptions for daily exercise, grocery shopping, and ‘essential’ trips; 3 - require not leaving house with minimal exceptions);
- Restrictions on internal movement: record restrictions on internal movement between cities/regions (0 - no measures; 1 - recommend not to travel between regions/cities; 2 - internal movement restrictions in place); and,
- International travel controls: record restrictions on international travel for foreign travelers (0 - no restrictions; 1 - screening arrivals; 2 - quarantine arrivals from some or all regions; 3 - ban arrivals from some regions; 4 - ban on all regions or total border closure). For each study included in the meta-analysis, we calculated the mean of the daily ordinal score of each of the above indexes, during two timeframes:
- 2-week: weeks before the start date of the study until the end date of the study; and,
- 4-week: weeks before the start date of the study until the end date of the study.

### 2.7. Statistical Analysis

We included all the rates (crude number cases/total number of individuals) in separate meta-analysis models for depression (PHQ-9 >= 10) and anxiety (GAD-7 >= 10). One study provided weighted rates for the outcomes only.^24^ We used a random-effects model because high heterogeneity was expected. We calculated I^2^ as a measure of between-study heterogeneity. Data were analyzed using OpenMetanalyst,^79^ which makes use of R metafor package.^80^ The threshold for significance was set to *p*-values of less than 0.05. In addition, we carried out further subgroup analysis models by population type (general, healthcare providers, students, patients, and mixed), region of the world (Asia, Europe, and Other), country (China and other), income level (high-income, and low-and middle-income), and non-national status (local studies were defined as those restricted to either a city or a state/province/region within a country, versus national studies).

Finally, we investigated the impact of physical distancing measures on depression and anxiety through meta-regression models.^81^ Separate models were carried out for different timeframes of physical distancing measures (2 and 4 weeks). Models adjusted for gender, sub-populations, timepoint when study began, region of the world, local status and previous levels of either depression or anxiety, depending on the outcome. Country indicators were not included in these models because of the strong correlation with earlier levels of depression and anxiety variables, which were collected based on previous data by each country. Meta-regression was used instead of subgroup analyses (i.e., different levels of social measures implementation) to allow for the use of continuous and multiple covariates. The random-effects meta-regression used residual restricted maximum likelihood to measure between-study variance (τ2) with a Knapp-Hartung modification as recommended models.^81^

## 3. Results

Table S1 presents the key-information of the 60 studies included. Eight studies were split into subsamples, and two studies reported the same sample. We included 67 samples in the meta-analysis models. All studies were conducted in 2020 (compiled date range of study initiation to closure: January 24^th^-May 31^st^), with a mean length of 15.4 days. In total, 226,638 individuals were included, with an average of 3,382 individuals per study. The mean age was 33.8 (range: 13-89) among samples that provided data on mean age and range, and the proportion of females included was 61.9% (range: 0-100). Few samples were representative (5.9%, N=4), and local (32.8%, N=22). Most samples were based in China (38.8%, N=26), and Asia in general (52.2%, N=35). General population samples were the most common (40.2%, N=27), followed by healthcare providers (23.8%, N=16), students (16.4%, N=11), and patients (8.9%, N=6). The vast majority of the samples used online methods (91.0%, N=61) and were peer-reviewed (64.1%, N=43). Table S2 presents the results of the quality assessment. All the included studies scored five or higher in such an assessment. Tables S3 and S4 present implementation of physical distancing measures and previous prevalence of depression and anxiety, respectively.

Figure 2 presents both the global results of the meta-analysis for depression and a subgroup analysis by region of the world (N=191,519). We found a global prevalence of 24.0% (95% Confidence Interval (CI): 21.0-27.1%) of depression; depression was observed among 17.6% (95%CI: 15.4-19.8%) in Asia, among 26.0% (95%CI: 22.9-29.05) in Europe, and among 39.1% (95%CI: 29.2-49.1%) in other regions of the world. A subgroup analysis (Figure S3) demonstrated that China had a lower prevalence of depression (16.2%, 95%CI:13.7-18.2%) than in other countries (29.0%, 95%CI:24.8-33.2). Additional subgroup analyses (Figures S4, S5, and S6) found no significant differences by population type, country income level, or being a local study.

Figure 3 presents the global results for anxiety, with a subgroup analysis by region of the world (N=193,137). We found a global prevalence of anxiety of 21.3% (95%CI:19.0-23.6%). Asia had lower levels of anxiety (17.9%, 95%CI:15.4-20.3) compared to other regions of the world (28.6%, 95%CI:22.6-34.6). Europe did not differ from Asia and the other regions of the world. Subgroup analysis at the country-level (Figure S7) showed that China had a lower prevalence of anxiety (15.5%, 95%CI:13.1-17.9%) compared to all other countries (25.6%, 95%CI:23.1-28.0). The number of studies in each of the other countries was too restrictive to make country-specific comparisons (i.e., U.S. was the second country with more studies having just 4 studies). Further subgroup analysis (Figures S8, S9, and S10) found no significant differences by population type, country income level, or being a local study.

Table 1 shows the results of the meta-regression models for depression. Both in the 2- and 4- week physical distancing models, previous depression, older studies, and other region of the world than Asia/Europe were associated with depression. In addition, patient studies had a higher prevalence of depression in the 2-week physical distancing model. No significant association with physical distancing implementation measures was found in both models.

Table 2 presents the results of the meta-regression models for anxiety. Both in the 2- and 4- week physical distancing models, the closure of public transportation was associated with anxiety. Student studies had lower levels of anxiety in both models. No other significant association between physical distancing measures and depression or anxiety were found.

## 4. Discussion

This study aimed to investigate levels of depression and anxiety during the COVID-19 pandemic and the effect of physical distancing measures on depression and anxiety. We found high global prevalences of both depression and anxiety during the COVID-19 pandemic (24.0% and 21.3%, respectively); however, there was a wide variance in the prevalence of both anxiety and depression reported at the region-and country-level. Asia, and China especially, presented lower levels of both anxiety and depression, compared to the other r and countries. Closure of public transportation increased levels of anxiety, independently of the timeframe (2 or 4 weeks post-transportation closure enactment).

As discussed by Galea et al.,^82^ the global healthcare sector must increase support for prevention and early intervention of depression and anxiety secondary to COVID-19 and physical distancing measures. Within the subgroup of Asian countries, estimates of depression prevalence ranged from 4.2-34.7%, with variance likely due to estimates derived from different Chinese provinces. When comparing to the prevalence of depression in the pre-COVID-19 era, ranging from 3.3-4.2%^83,84^, these estimates are demonstrably larger after the initiation of COVID-19. This pattern is upheld for the remaining countries classified within the Asian region.

Prior to the occurrence of COVID-19, the prevalence of depression reported in: Korea was estimated to be 6.1%;^85^ Pakistan was 4.5%;^83^ Nepal was 16.8%;^86^ and Japan was 7.9%.^87^ Similarly, pre-COVID rates of depression within the subgroup of countries classified as Other ranged from: 3.9% in Nigeria;^83^ 4.0% in Jordan;^83^ 4.9% in Iran;^83^ 4.5% in Saudi Arabia;^83^ 4.2-4.6% in Brazil;^88^ and 8% in the United States.^89^ Within the European countries, reported prevalences of depression prior to COVID-19 include: 4.5% in the UK;^83^ 4.8% in Albania;^83^ 3.6-5% in Switzerland;^83,90^ 5.1% in Italy and Austria;^83^ 5.2% in Spain;^83^ 2.6-8.5% in Norway;^91^ and 6.1-10.2% in Germany.^92^ The only country to report potentially lower depression rates post-COVID-19 is Russia; however, pre-COVID-19 estimates range from 5.5%^83^ to 31.2-37.8%,^93^ a variation which may be the result of differing scales or methods for assessing depression. Overall, we observe a marked increase in depression prevalence in the post-COVID-19 era.

Similarly, the prevalence of anxiety, as reported in the subgroup of Asian countries is largely greater subsequent to the onset of COVID-19. Rates of anxiety prior to COVID-19 ranged from: 3.0-5.8% in India;^83,94^ 3.1% in China;^83^ 3.2% in Nepal;^83^ 4.1% in Bangladesh;^83^ and 4.2% in Pakistan.^83^ Japan, however, reported a prevalence of anxiety of 22.6%,^95^ which is higher than the prevalence of 10.9% reported post-COVID-19.^56^ Increases in anxiety can further be observed in the countries classified within the Other category. Prior to COVID-19, anxiety rates were: 2.7% in Nigeria;^83^ 4.1% in the United Arab Emirates;^83^ 4.3% in Saudi Arabia and Jordan;^83^ 12.1-12.7% in Brazil;^88^ and 8.2% in the United States.^96^ Among the European countries, estimates of anxiety prevalence prior to COVID were: 3.8% in Serbia;^83^ 4.9% in Switzerland;^83^ 5% in Italy;^83^ 5.1% in Cyprus;^83^ 6.5% in Austria;^97^ 6.6% in Norway;^98^ 7.2% in the United Kingdom;^99^ and 9.7% in Spain.^100^ Russia and Germany both reported higher anxiety prevalences of 22.0%^93^ and 19.0%^101^, respectively, in comparison to rates observed subsequent to the occurrence of COVID-19.

Our finding regarding the effect of public transportation closures on anxiety levels points to the importance of these systems to global populations. We understand that anxiety could emerge as a result of two fear/worry dimensions: not being able to achieve the basic needs and/or insecurity. Depending on the setting (i.e., rural, small to large metropolitan areas), there is a significant number of individuals who do not have an alternative way of transport (i.e., car, motorcycle) and are dependent on public transportation.^102^ People in many different countries and cultural contexts rely on some method of public transport for getting food, clothing, education, shelter, healthcare, sanitation,^103^ such as transport within metropolitan areas to places of employment.^104^ It is thus reasonable to theorize that anxious anticipatory thinking could emerge in people dependent on public transport. These are core symptoms of many anxiety disorders,^105^ which are captured by our anxiety outcome measure (GAD-7). In addition to worry regarding reliability of public transport, anxiety could grow from increased risk of assault and harassment resultant from fewer bystanders accessing this method of transportation.^106^ Considering that mitigation strategies in the COVID era have involved significantly reducing the volume of passengers, the number of routes, and the means of transport available,^107^ closures of these systems can work to generate excessive anxiety and worry.^108^

### 4.1. Strengths and Limitations

Country-level data of physical distancing measures and previous anxiety or depression is an important limitation of the present study. However, we included data from 67 different samples from 26 countries, within five global regions (Asia, Africa, America, Europe, and Middle-East), totaling almost 200,000 individuals in each meta-analysis. In addition, we used just one outcome measure per disorder (PHQ-9 and GAD-7), to avoid outcome measure bias, common in meta-analysis studies. Unfortunately, we were not able to include age as a covariate in the meta-regression models due to lack of descriptive data. A portion of the included samples (35.9%, N=24) were not peer-reviewed. Notably, inclusion of data from pre-print repositories could be seen as both a strength and limitation, in that the inclusion of the most recent data is of utmost importance. Results should be interpreted with caution.

### 4.2. Conclusion

The COVID-19 pandemic, and the resulting physical distancing measures to mitigate viral spread, has impacted population mental health worldwide. Despite finding a wide variation in anxiety and depression levels across countries and regions of the world, high prevalence of mental health disorders is a considerable concern during the COVID era. Thus, mental health outcomes should not be addressed as a delayed consequence of the COVID-19 pandemic, but rather as an ongoing and concurrent epidemic (i.e., a syndemic). We also observed an association between restrictions and closures of public transportation systems and an increase in anxiety levels. These results have important implications for policymakers. There is an urgent need for the healthcare sector to increase now support for prevention and early intervention of depression and anxiety.

## Data Availability

All the data used is presented in the manuscript.

---

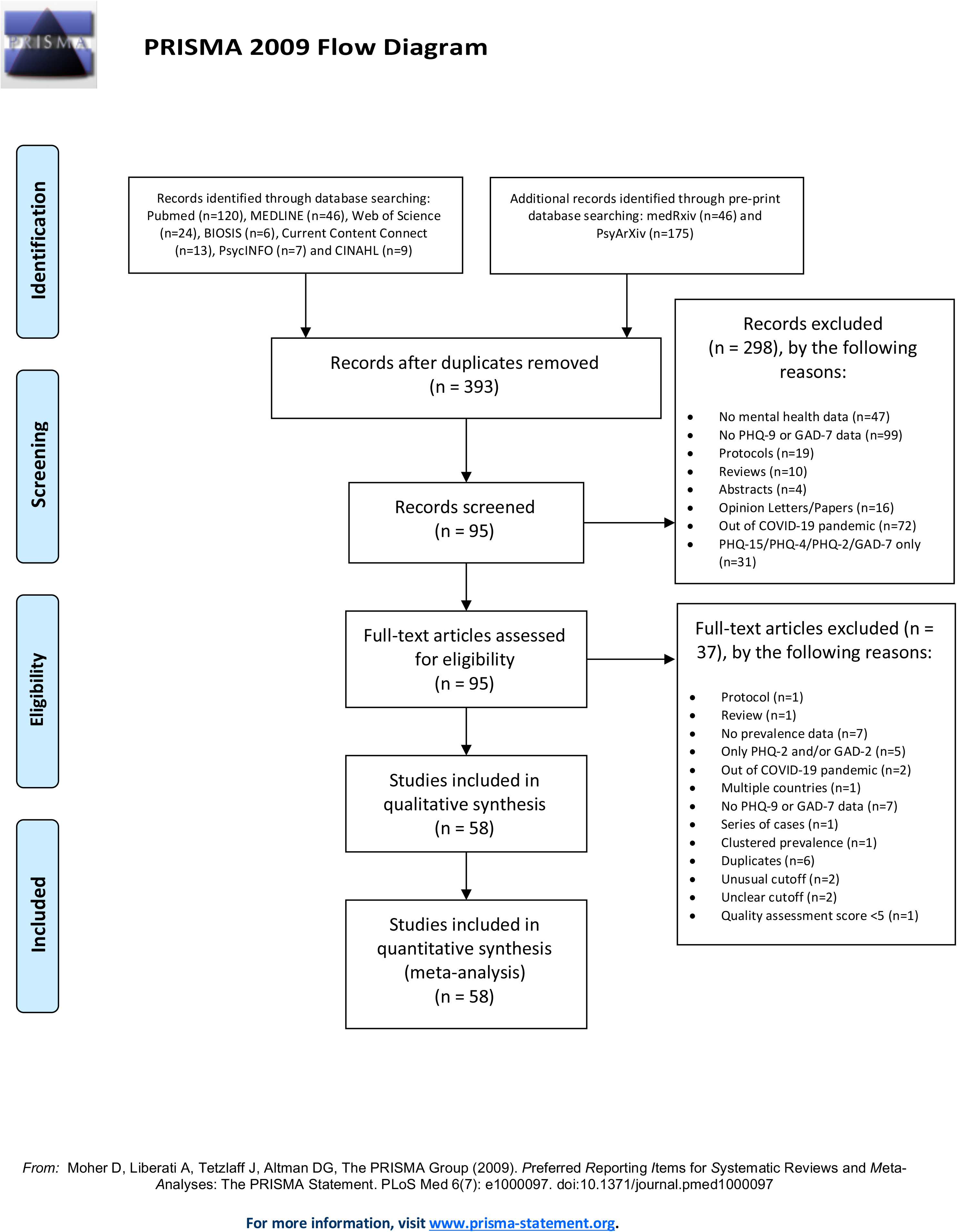

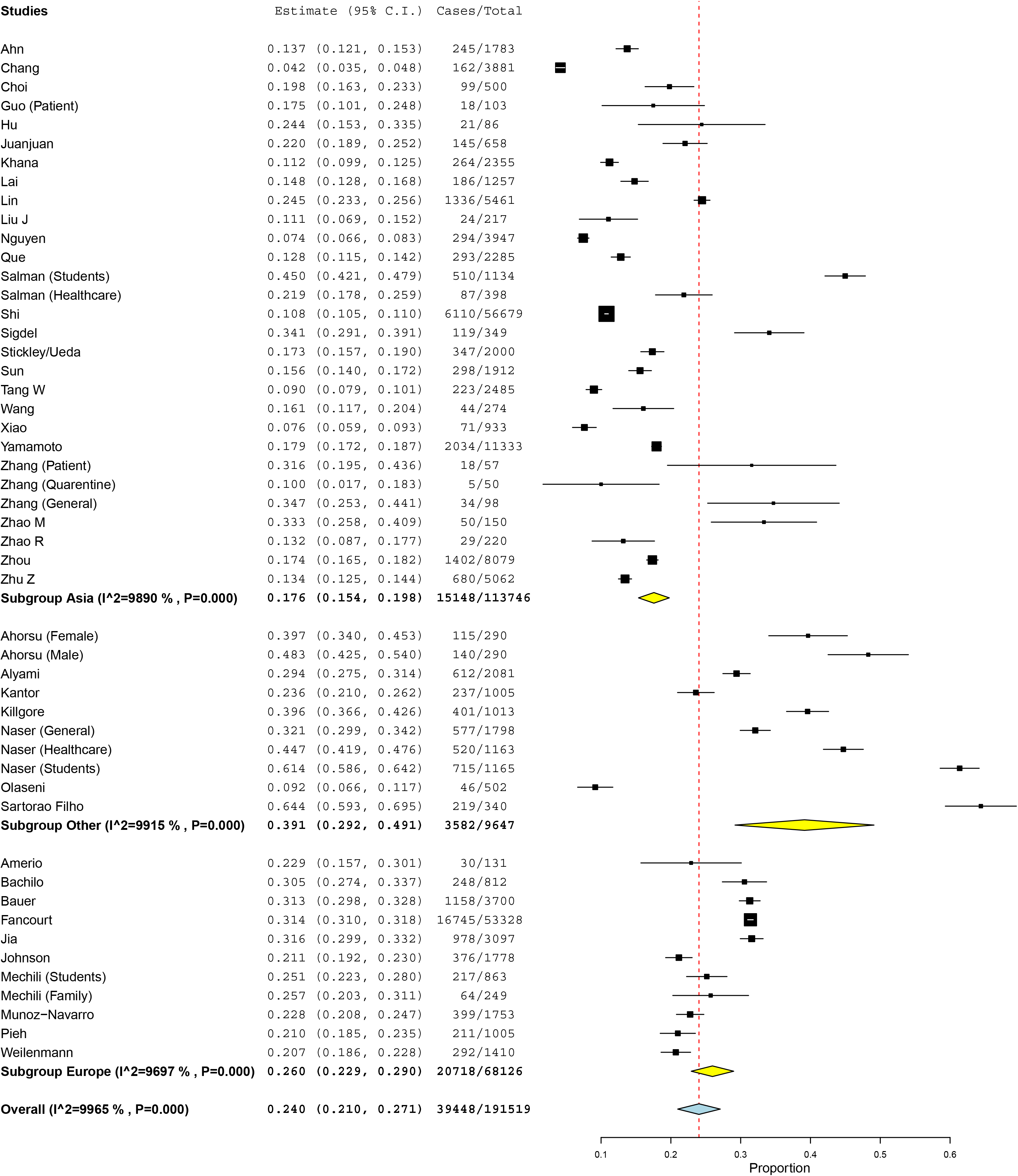
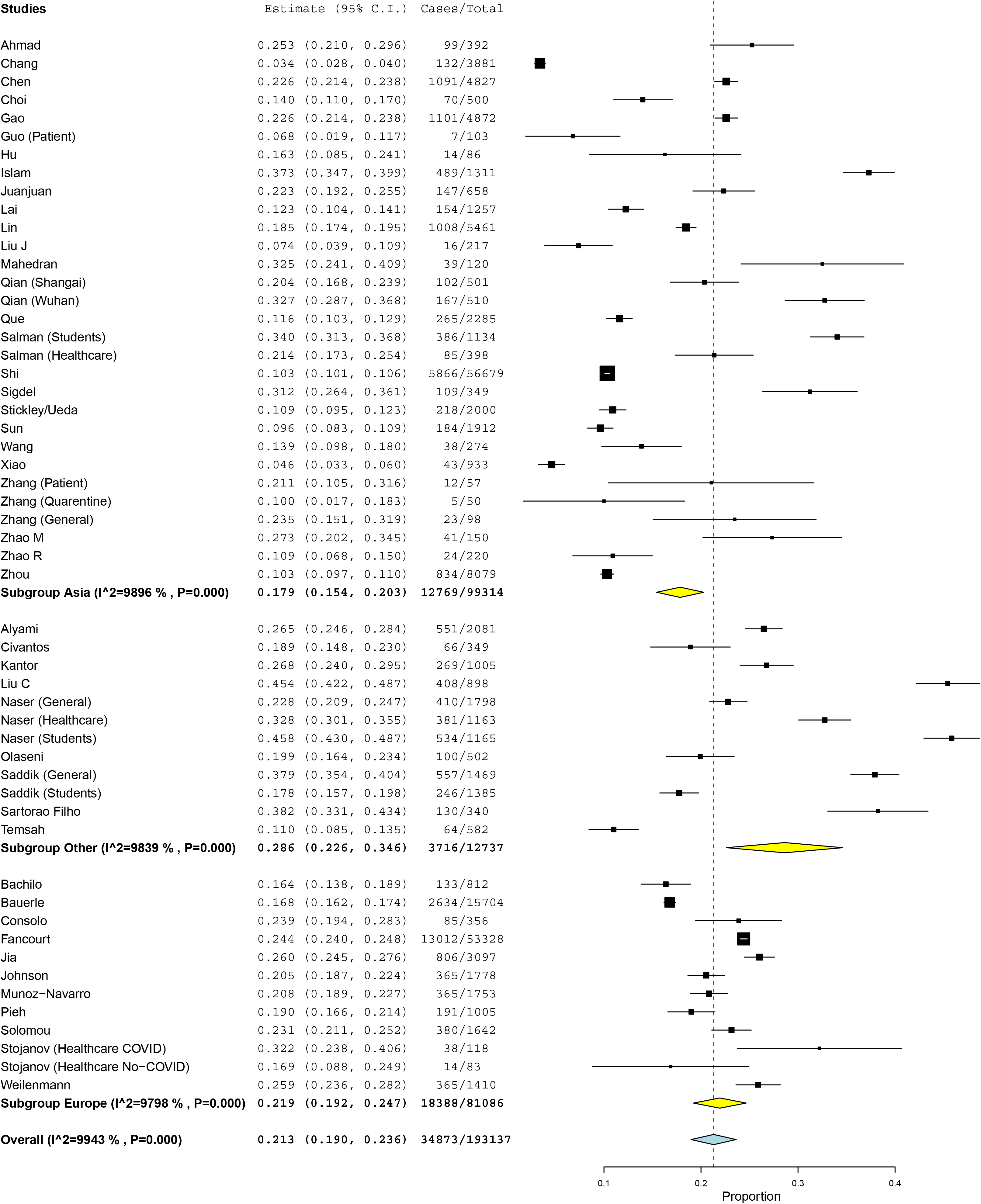

**Table 1.**
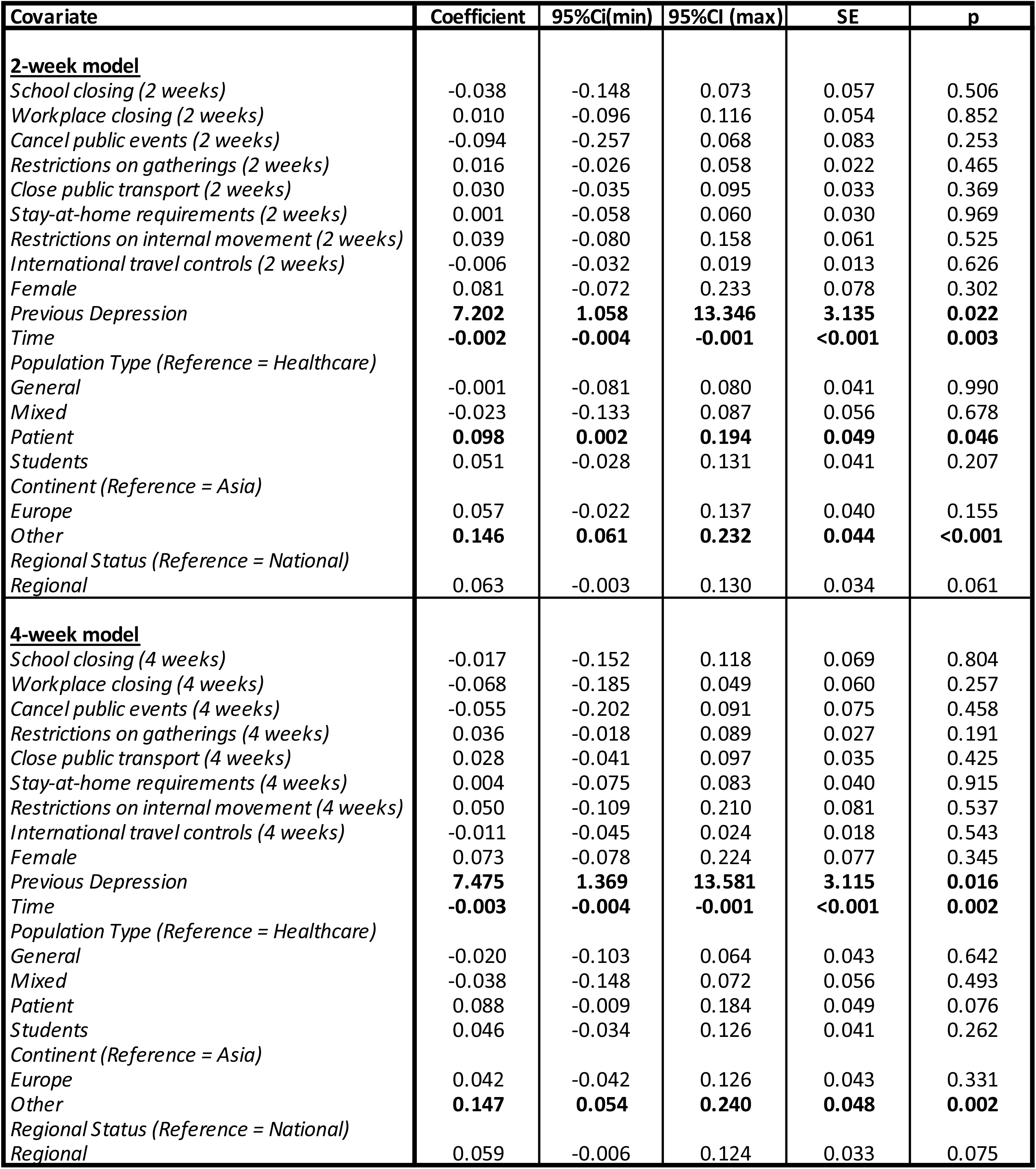
Results for the meta-regression models for depression.

**Table 2.** Results for the meta-regression models for anxiety.

| Covariate | Coefficient | 95%CI (min) | 95%CI (max) | SE | p |
| --- | --- | --- | --- | --- | --- |
| **<u>2-week model</u>** |  |  |  |  |  |
| *School closing (2 weeks)* | 0.002 | -0.093 | 0.098 | 0.049 | 0.961 |
| *Workplace closing (2 weeks)* | -0.001 | -0.099 | 0.097 | 0.050 | 0.988 |
| *Cancel public events (2 weeks)* | 0.007 | -0.195 | 0.208 | 0.103 | 0.949 |
| *Restrictions on gatherings (2 weeks)* | 0.022 | -0.065 | 0.109 | 0.044 | 0.615 |
| *Close public transport (2 weeks)* | **0.071** | **0.007** | **0.134** | **0.032** | **0.029** |
| *Stay-at-home requirements (2 weeks)* | -0.029 | -0.096 | 0.038 | 0.034 | 0.399 |
| *Restrictions on internal movement (2 weeks)* | -0.043 | -0.173 | 0.086 | 0.066 | 0.512 |
| *International travel controls (2 weeks)* | 0.005 | -0.019 | 0.030 | 0.013 | 0.676 |
| *Female* | 0.080 | -0.093 | 0.253 | 0.088 | 0.366 |
| *Previous Anxiety* | 1.408 | -2.472 | 5.288 | 1.980 | 0.477 |
| *Time* | -0.001 | -0.002 | 0.001 | <0.001 | 0.286 |
| *Population Type (Reference = General)* |  |  |  |  |  |
| *Healthcare* | -0.040 | -0.107 | 0.026 | 0.034 | 0.237 |
| *Mixed* | -0.039 | -0.127 | 0.049 | 0.045 | 0.385 |
| *Patient* | -0.017 | -0.104 | 0.070 | 0.044 | 0.706 |
| *Students* | **-0.068** | **-0.133** | **-0.003** | **0.033** | **0.041** |
| *Continent (Reference = Asia)* |  |  |  |  |  |
| *Europe* | 0.003 | -0.100 | 0.106 | 0.053 | 0.955 |
| *Other* | 0.091 | -0.017 | 0.199 | 0.055 | 0.100 |
| *Regional Status (Reference = National)* |  |  |  |  |  |
| *Regional* | 0.017 | -0.036 | 0.071 | 0.027 | 0.530 |
| **<u>4-week model</u>** |  |  |  |  |  |
| *School closing (4 weeks)* | -0.060 | -0.162 | 0.043 | 0.052 | 0.256 |
| *Workplace closing (4 weeks)* | -0.014 | -0.119 | 0.091 | 0.054 | 0.792 |
| *Cancel public events (4 weeks)* | 0.041 | -0.095 | 0.177 | 0.069 | 0.551 |
| *Restrictions on gatherings (4 weeks)* | 0.032 | -0.038 | 0.101 | 0.035 | 0.368 |
| *Close public transport (4 weeks)* | **0.066** | **0.004** | **0.128** | **0.032** | **0.038** |
| *Stay-at-home requirements (4 weeks)* | -0.031 | -0.104 | 0.043 | 0.038 | 0.414 |
| *Restrictions on internal movement (4 weeks)* | -0.030 | -0.166 | 0.106 | 0.069 | 0.669 |
| *International travel controls (4 weeks)* | 0.017 | -0.013 | 0.048 | 0.016 | 0.267 |
| *Female* | 0.025 | -0.143 | 0.193 | 0.086 | 0.769 |
| *Previous Anxiety* | 1.442 | -1.906 | 4.789 | 1.708 | 0.399 |
| *Time* | -0.001 | -0.003 | 0.000 | <0.001 | 0.060 |
| *Population Type (Reference = General)* |  |  |  |  |  |
| *Healthcare* | -0.059 | -0.122 | 0.004 | 0.032 | 0.067 |
| *Mixed* | -0.031 | -0.108 | 0.046 | 0.039 | 0.426 |
| *Patient* | -0.005 | -0.087 | 0.078 | 0.042 | 0.911 |
| *Students* | **-0.068** | **-0.130** | **-0.006** | **0.032** | **0.033** |
| *Continent (Reference = Asia)* |  |  |  |  |  |
| *Europe* | -0.022 | -0.115 | 0.072 | 0.048 | 0.649 |
| *Other* | 0.061 | -0.028 | 0.151 | 0.046 | 0.179 |
| *Regional Status (Reference = National)* |  |  |  |  |  |
| *Regional* | -0.008 | -0.060 | 0.044 | 0.026 | 0.766 |

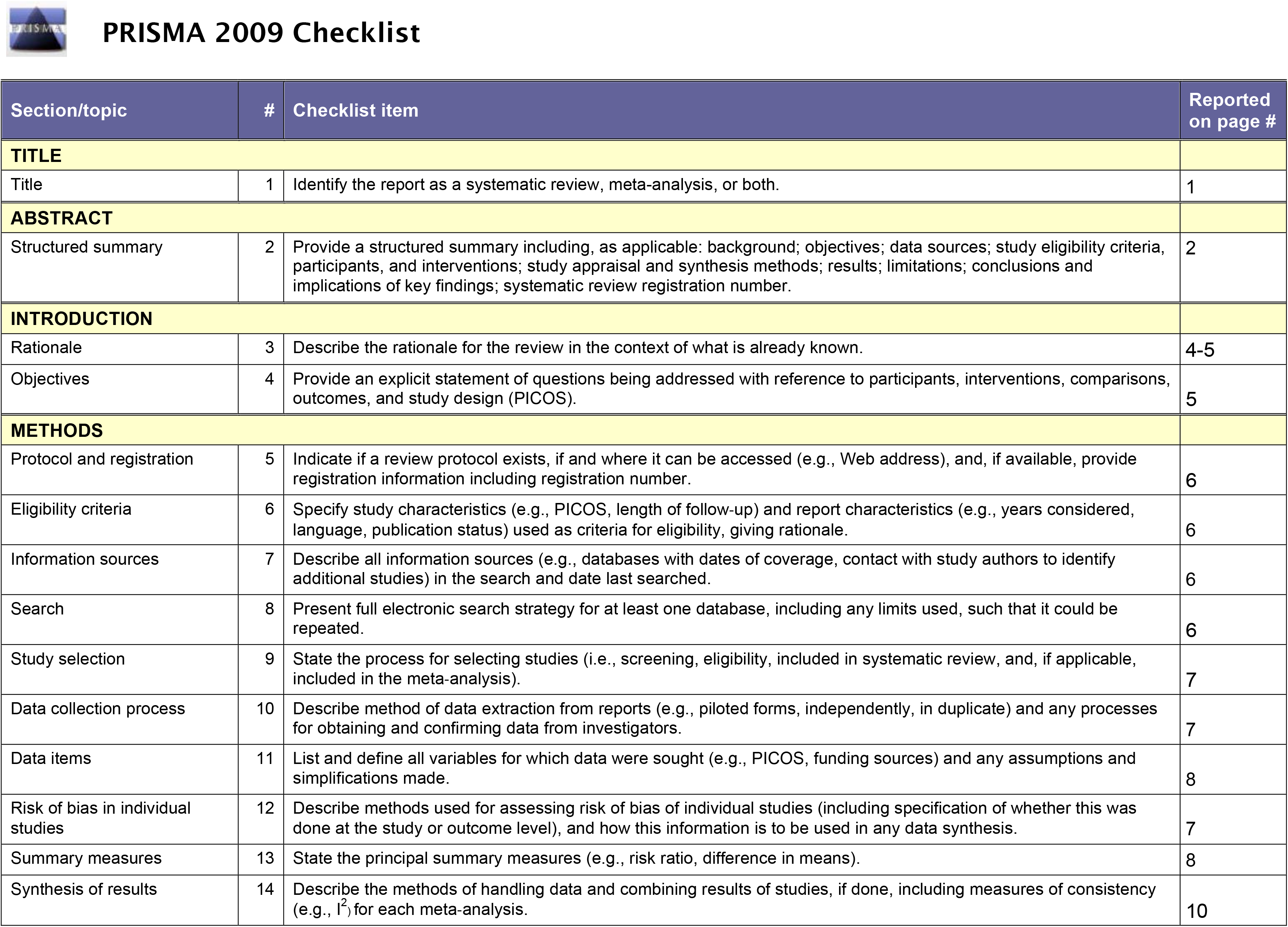
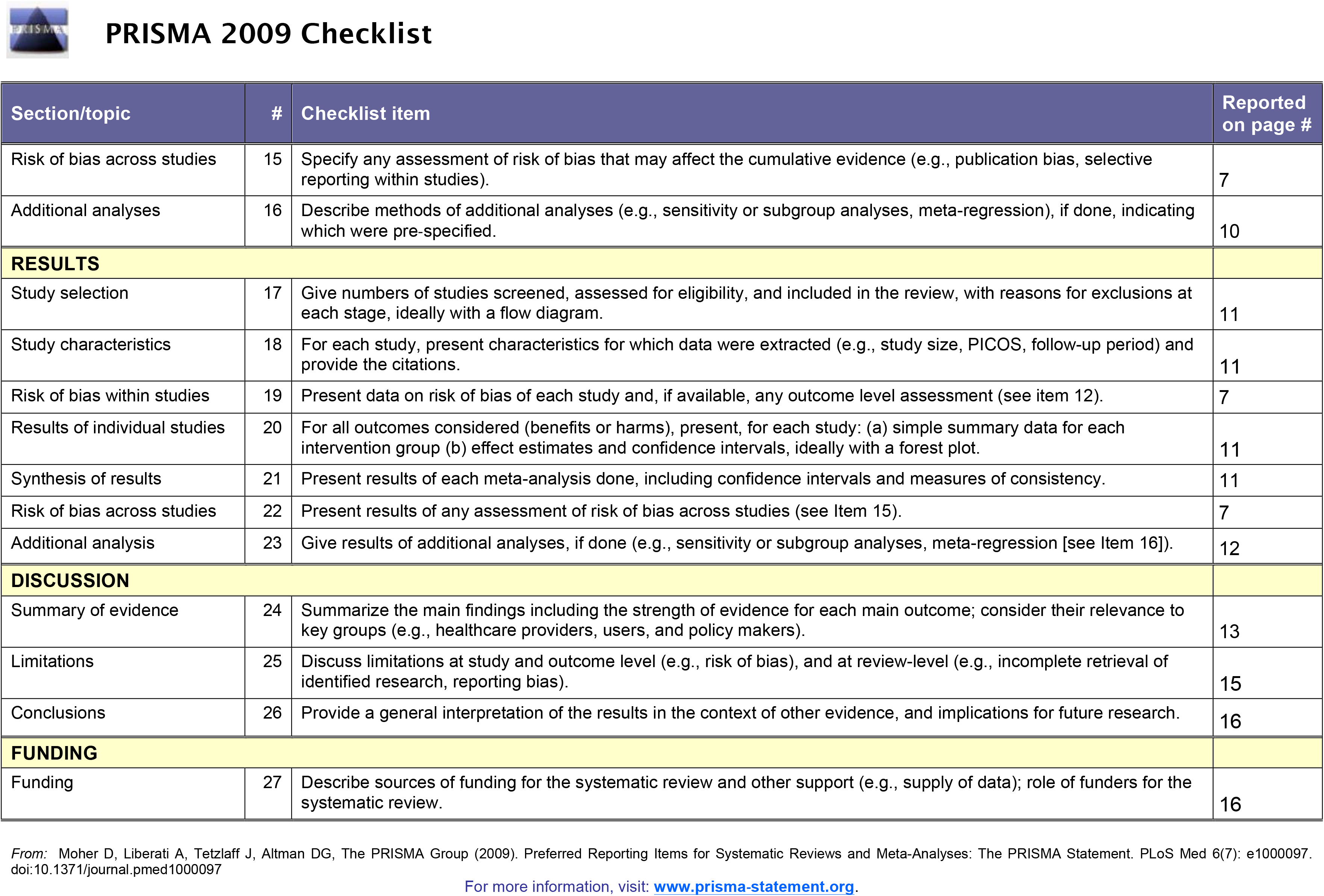

## MOOSE (Meta-analyses Of Observational Studies in Epidemiology) Checklist

A reporting checklist for Authors, Editors, and Reviewers of Meta-analyses of Observational Studies. You must report the page number in your manuscript where you consider each of the items listed in this checklist. If you have not included this information, either revise your manuscript accordingly before submitting or note N/A.

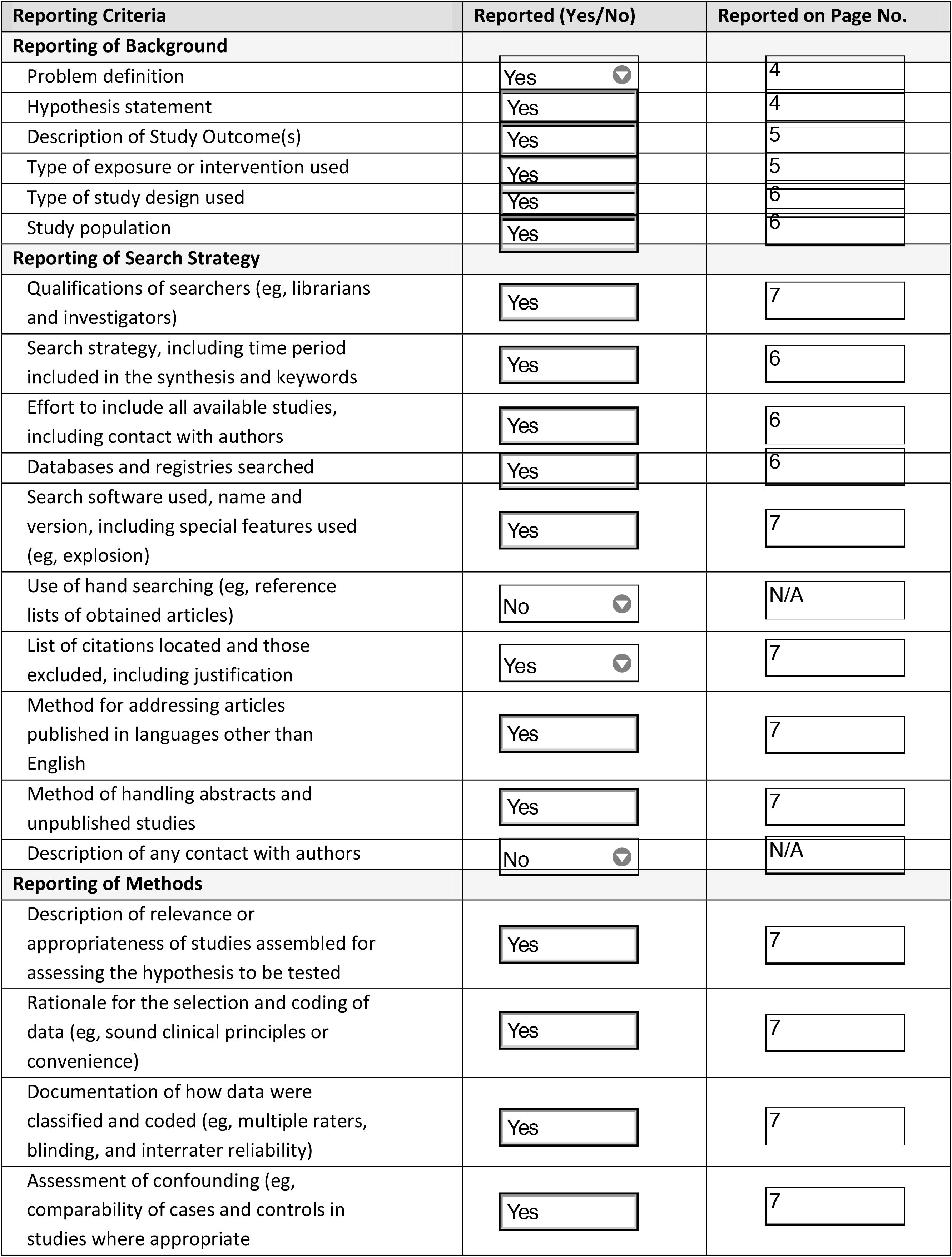
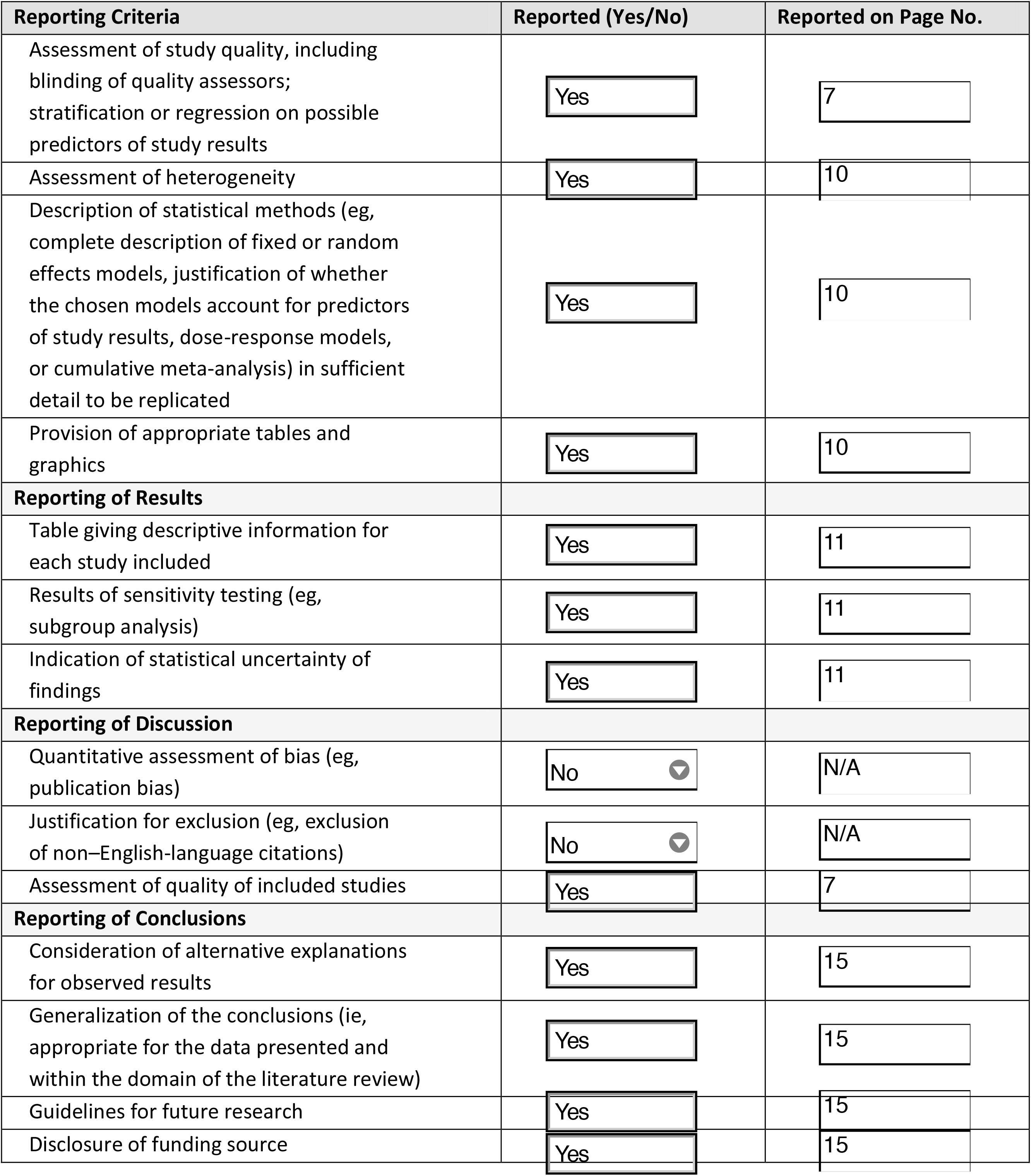

**Once you have completed this checklist, please save a copy and upload it as part of your submission. DO NOT include this checklist as part of the main manuscript document. It must be uploaded as a separate file**.

**Table S1.**
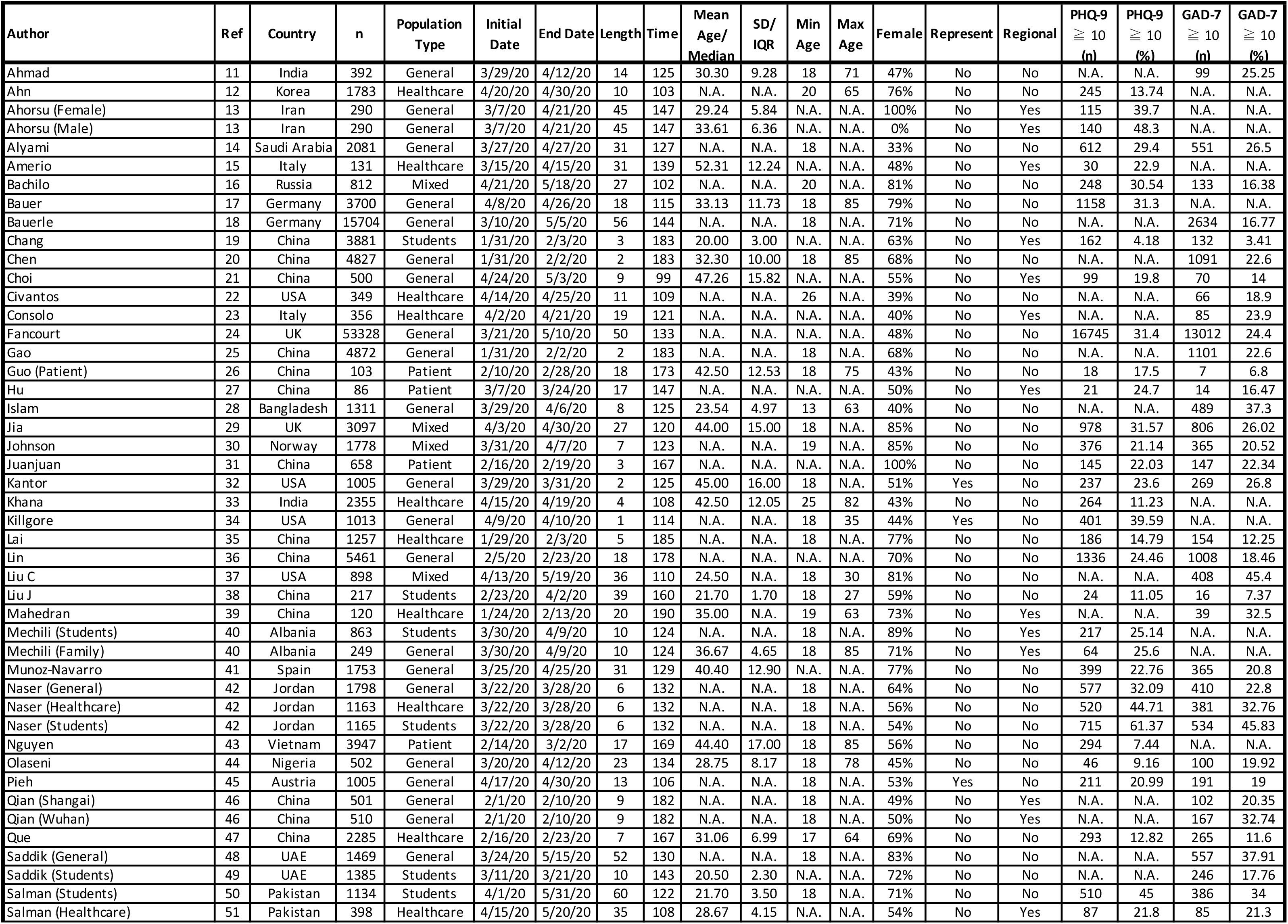
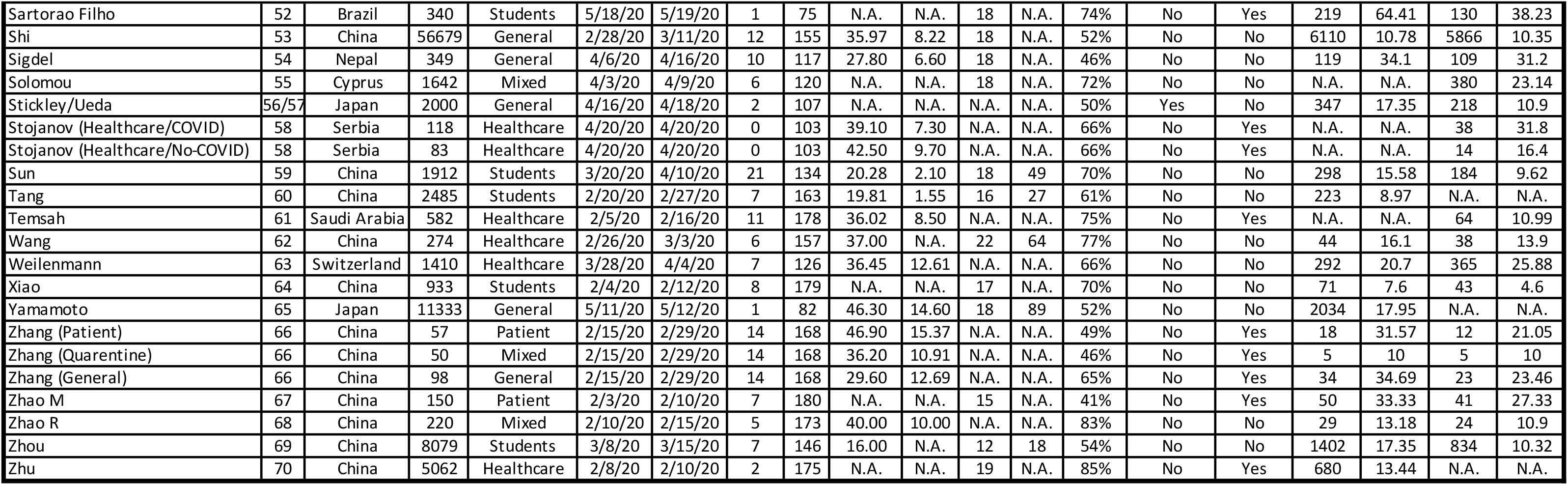
Main findings from the included studies.

**Table S2.**
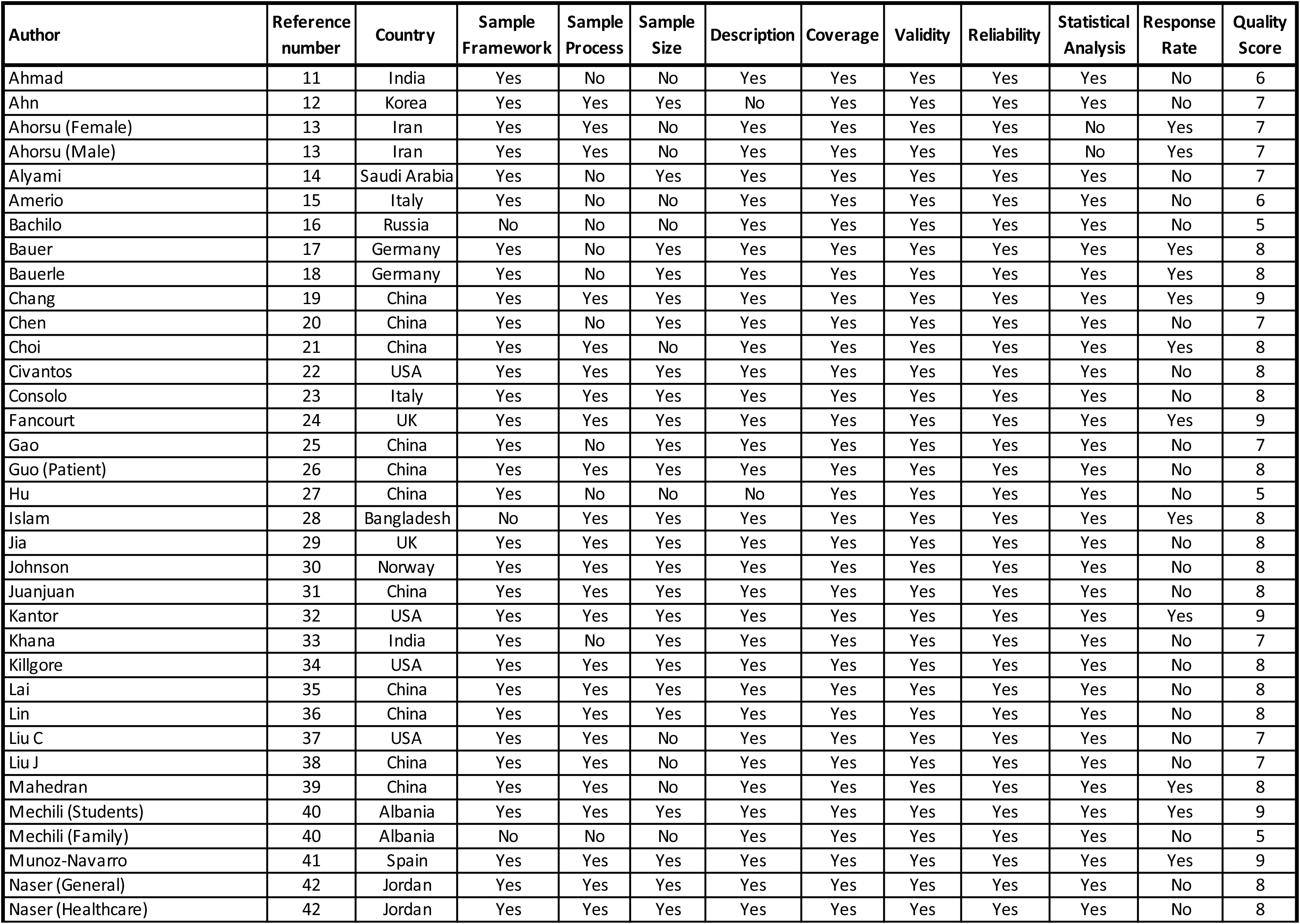
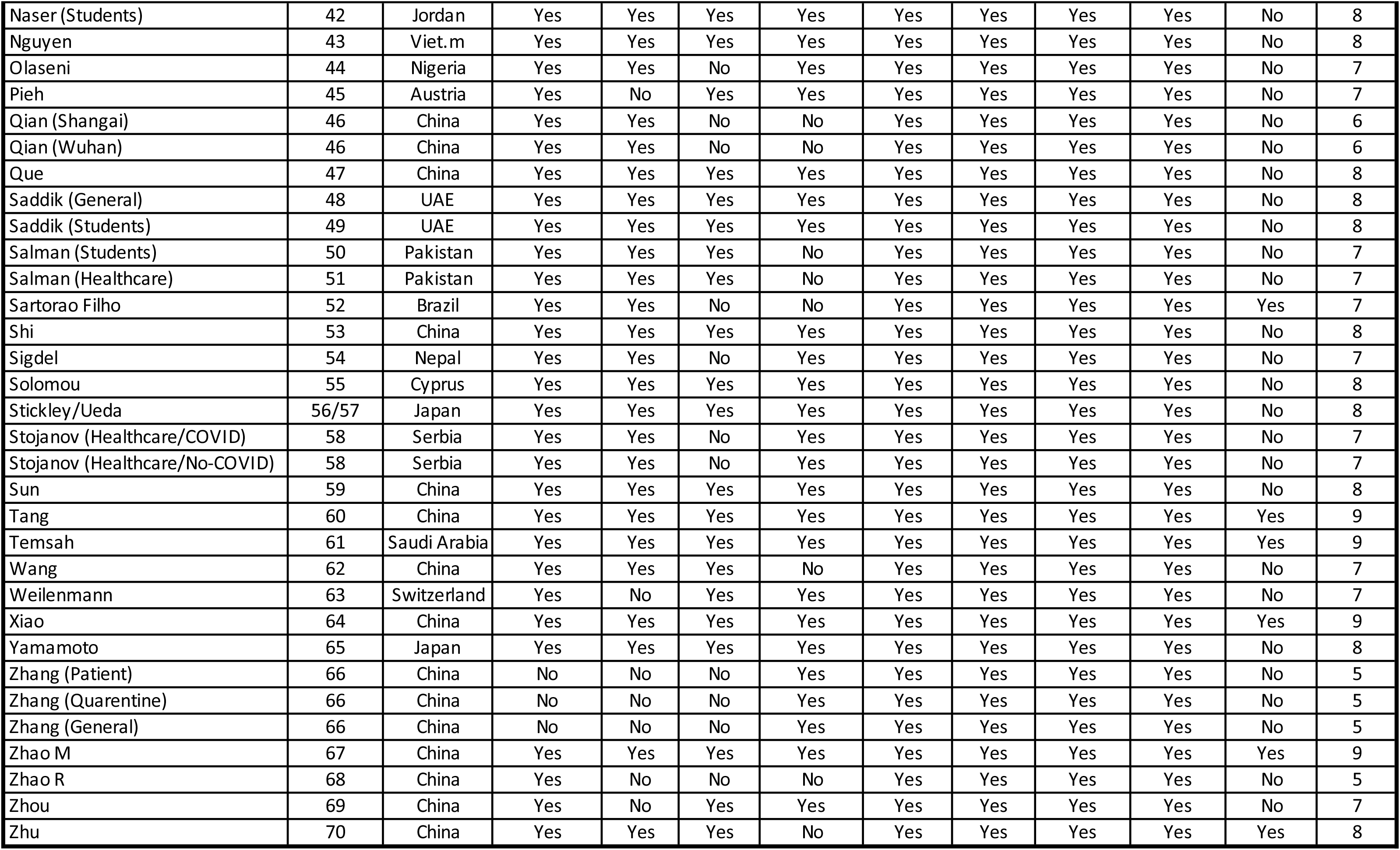
Quality Assessment Results.

**Table S3.** Mean of social isolation measures implementation national data based on Oxford Covid-19 Government Response Tracker (Hale et al., 2020), during the period of each study.

| Author | Ref | Country | School closing (2 weeks) | Workplace closing (2 weeks) | Cancel public events (2 weeks) | Restrict. on gather. (2 weeks) | Close public transp. (2 weeks) | Stay-at-home requir. (2 weeks) | Restrict. on internal mov. (2 weeks) | Intern. travel controls (2 weeks) | School closing (4 weeks) | Workplace closing (4 weeks) | Cancel public events (4 weeks) | Restrict. on gather. (4 weeks) | Close public transp. (4 weeks) | Stay-at-home requir. (4 weeks) | Restrict. on internal mov. (4 weeks) | Intern. travel controls (4 weeks) |
| --- | --- | --- | --- | --- | --- | --- | --- | --- | --- | --- | --- | --- | --- | --- | --- | --- | --- | --- |
| Ahmad | 11 | India | 3.000 | 2.552 | 2.000 | 3.448 | 1.655 | 2.517 | 1.793 | 3.759 | 2.581 | 1.721 | 1.605 | 2.326 | 1.116 | 2.023 | 1.209 | 2.907 |
| Ahn | 12 | Korea | 3.000 | 2.483 | 1.621 | 2.414 | 0.000 | 1.172 | 1.172 | 3.000 | 2.997 | 2.356 | 1.728 | 2.591 | 0.073 | 1.433 | 1.396 | 3.025 |
| Ahorsu (Female) | 13 | Iran | 2.441 | 1.814 | 2.000 | 0.051 | 0.712 | 0.576 | 1.627 | 0.000 | 1.973 | 1.466 | 1.671 | 0.041 | 0.575 | 0.466 | 1.315 | 0.000 |
| Ahorsu (Male) | 13 | Iran | 2.441 | 1.814 | 2.000 | 0.051 | 0.712 | 0.576 | 1.627 | 0.000 | 1.973 | 1.466 | 1.671 | 0.041 | 0.575 | 0.466 | 1.315 | 0.000 |
| Alyami | 14 | Saudi Arabia | 3.000 | 1.458 | 1.559 | 2.220 | 1.288 | 1.593 | 1.729 | 2.983 | 2.500 | 1.506 | 1.599 | 1.889 | 1.210 | 1.459 | 1.724 | 2.479 |
| Amerio | 15 | Italy | 3.000 | 3.000 | 2.000 | 4.000 | 0.833 | 2.438 | 2.000 | 3.000 | 2.565 | 2.613 | 1.710 | 3.419 | 0.645 | 2.048 | 1.774 | 3.000 |
| Bachilo | 16 | Russia | 3.000 | 2.833 | 2.000 | 4.000 | 1.000 | 3.000 | 2.000 | 4.000 | 3.000 | 2.768 | 2.000 | 3.893 | 0.893 | 2.893 | 2.000 | 3.893 |
| Bauer | 17 | Germany | 3.000 | 2.000 | 2.000 | 4.000 | 0.000 | 2.000 | 2.000 | 4.000 | 2.894 | 1.532 | 2.000 | 3.362 | 0.000 | 1.787 | 1.681 | 3.638 |
| Bauerle | 18 | Germany | 2.662 | 1.268 | 1.746 | 2.746 | 0.000 | 1.465 | 1.366 | 3.085 | 2.224 | 1.059 | 1.459 | 2.294 | 0.000 | 1.224 | 1.141 | 2.576 |
| Chang | 19 | China | 1.500 | 1.500 | 1.444 | 2.889 | 1.333 | 1.000 | 1.333 | 0.000 | 0.844 | 0.844 | 0.813 | 1.625 | 0.750 | 0.563 | 0.750 | 0.000 |
| Chen | 20 | China | 1.412 | 1.412 | 1.412 | 2.824 | 1.294 | 0.882 | 1.294 | 0.000 | 0.774 | 0.774 | 0.774 | 1.548 | 0.710 | 0.484 | 0.710 | 0.000 |
| Choi | 21 | China | 3.000 | 2.000 | 2.000 | 4.000 | 0.000 | 1.000 | 1.000 | 3.000 | 3.000 | 2.184 | 2.000 | 4.000 | 0.053 | 1.632 | 1.316 | 3.000 |
| Civantos | 22 | USA | 3.000 | 3.000 | 2.000 | 4.000 | 1.000 | 2.000 | 2.000 | 3.000 | 3.000 | 2.850 | 2.000 | 3.800 | 1.000 | 2.000 | 1.950 | 3.000 |
| Consolo | 23 | Italy | 3.000 | 3.000 | 2.000 | 4.000 | 1.286 | 2.600 | 2.000 | 3.000 | 3.000 | 3.000 | 2.000 | 4.000 | 1.061 | 2.429 | 2.000 | 3.000 |
| Fancourt | 24 | UK | 2.262 | 1.646 | 1.631 | 3.015 | 0.708 | 1.508 | 1.523 | 0.000 | 1.861 | 1.354 | 1.342 | 2.481 | 0.582 | 1.241 | 1.253 | 0.000 |
| Gao | 25 | China | 1.263 | 1.263 | 1.263 | 2.526 | 1.158 | 0.789 | 1.158 | 0.000 | 0.774 | 0.774 | 0.774 | 1.548 | 0.710 | 0.484 | 0.710 | 0.000 |
| Guo (Patient) | 26 | China | 3.000 | 3.000 | 2.000 | 4.000 | 2.000 | 2.697 | 2.000 | 0.242 | 2.170 | 2.170 | 1.617 | 3.234 | 1.574 | 1.979 | 1.574 | 0.170 |
| Hu | 27 | China | 3.000 | 3.000 | 2.000 | 4.000 | 2.000 | 3.000 | 2.000 | 1.813 | 3.000 | 3.000 | 2.000 | 4.000 | 2.000 | 3.000 | 2.000 | 1.261 |
| Islam | 28 | Bangladesh | 2.870 | 2.478 | 1.696 | 3.304 | 1.652 | 1.087 | 1.652 | 2.261 | 1.784 | 1.541 | 1.054 | 2.054 | 1.027 | 0.676 | 1.027 | 1.784 |
| Jia | 29 | UK | 2.786 | 1.976 | 1.976 | 3.714 | 0.857 | 1.857 | 1.881 | 0.000 | 2.089 | 1.554 | 1.536 | 2.786 | 0.643 | 1.393 | 1.411 | 0.000 |
| Johnson | 30 | Norway | 3.000 | 2.000 | 1.364 | 3.364 | 1.000 | 0.000 | 2.000 | 4.000 | 2.250 | 1.556 | 0.833 | 2.389 | 0.750 | 0.000 | 1.278 | 2.722 |
| Juanjuan | 31 | China | 3.000 | 3.000 | 2.000 | 4.000 | 2.000 | 3.000 | 2.000 | 0.000 | 2.344 | 2.344 | 1.813 | 3.625 | 1.750 | 2.063 | 1.750 | 0.000 |
| Kantor | 32 | USA | 3.000 | 2.294 | 2.000 | 3.294 | 0.882 | 2.000 | 1.765 | 3.000 | 2.613 | 1.258 | 1.645 | 2.032 | 0.484 | 1.097 | 1.000 | 2.968 |
| Kha. | 33 | India | 3.000 | 3.000 | 2.000 | 4.000 | 2.000 | 3.000 | 2.000 | 4.000 | 3.000 | 2.818 | 2.000 | 3.879 | 1.879 | 2.758 | 1.939 | 3.879 |
| Killgore | 34 | USA | 3.000 | 3.000 | 2.000 | 4.000 | 1.000 | 2.000 | 2.000 | 3.000 | 3.000 | 2.300 | 2.000 | 3.400 | 0.833 | 1.800 | 1.700 | 3.000 |
| Lai | 35 | China | 1.350 | 1.350 | 1.300 | 2.600 | 1.200 | 0.900 | 1.200 | 0.000 | 0.794 | 0.794 | 0.765 | 1.529 | 0.706 | 0.529 | 0.706 | 0.000 |
| Lin | 36 | China | 2.636 | 2.636 | 2.000 | 4.000 | 1.939 | 2.364 | 1.939 | 0.000 | 1.851 | 1.851 | 1.404 | 2.809 | 1.362 | 1.660 | 1.362 | 0.000 |
| Liu C | 37 | USA | 3.000 | 3.000 | 2.000 | 4.000 | 1.000 | 2.000 | 2.000 | 3.000 | 3.000 | 2.862 | 2.000 | 3.846 | 0.985 | 2.000 | 1.954 | 3.000 |
| Liu J | 38 | China | 3.000 | 3.000 | 2.000 | 4.000 | 1.778 | 3.000 | 2.000 | 1.556 | 3.000 | 3.000 | 2.000 | 4.000 | 1.824 | 2.824 | 2.000 | 1.235 |
| Mahedran | 39 | China | 1.629 | 1.629 | 1.314 | 2.629 | 1.257 | 1.371 | 1.257 | 0.000 | N.A. | N.A. | N.A. | N.A. | N.A. | N.A. | N.A. | N.A. |
| Mechili (Students) | 40 | Albania | 3.000 | 2.000 | 2.000 | 2.000 | 2.000 | 2.000 | 2.000 | 3.692 | 2.462 | 1.538 | 1.641 | 1.641 | 1.436 | 1.436 | 1.436 | 2.897 |
| Mechili (Family) | 40 | Albania | 3.000 | 2.000 | 2.000 | 2.000 | 2.000 | 2.000 | 2.000 | 3.692 | 2.462 | 1.538 | 1.641 | 1.641 | 1.436 | 1.436 | 1.436 | 2.897 |
| Munoz-varro | 41 | Spain | 3.000 | 2.522 | 2.000 | 2.761 | 0.935 | 1.870 | 1.000 | 3.870 | 2.400 | 1.967 | 1.567 | 2.133 | 0.717 | 1.433 | 0.800 | 3.017 |
| Naser (General) | 42 | Jordan | 2.000 | 1.571 | 1.048 | 2.095 | 1.048 | 1.571 | 1.048 | 2.667 | 1.200 | 0.943 | 0.629 | 1.257 | 0.629 | 0.943 | 0.629 | 1.600 |
| Naser (Healthcare) | 42 | Jordan | 2.000 | 1.571 | 1.048 | 2.095 | 1.048 | 1.571 | 1.048 | 2.667 | 1.200 | 0.943 | 0.629 | 1.257 | 0.629 | 0.943 | 0.629 | 1.600 |
| Naser (Students) | 42 | Jordan | 2.000 | 1.571 | 1.048 | 2.095 | 1.048 | 1.571 | 1.048 | 2.667 | 1.200 | 0.943 | 0.629 | 1.257 | 0.629 | 0.943 | 0.629 | 1.600 |
| Nguyen | 43 | Viet.m | 1.737 | 1.474 | 1.316 | 2.474 | 0.789 | 0.947 | 0.789 | 2.211 | 1.269 | 1.077 | 0.962 | 1.808 | 0.577 | 0.692 | 0.577 | 1.615 |
| Olaseni | 44 | Nigeria | 3.000 | 2.292 | 2.000 | 4.000 | 1.000 | 2.000 | 1.667 | 3.000 | 3.000 | 2.553 | 2.000 | 4.000 | 1.000 | 2.000 | 1.789 | 3.000 |
| Pieh | 45 | Austria | 2.000 | 2.000 | 1.667 | 3.333 | 1.583 | 1.625 | 1.583 | 0.000 | 1.263 | 1.263 | 1.053 | 2.105 | 1.000 | 1.026 | 1.000 | 0.000 |
| Qian (Shangai) | 46 | China | 3.000 | 3.000 | 2.000 | 4.000 | 2.000 | 3.000 | 2.000 | 0.000 | 2.417 | 2.417 | 1.833 | 3.667 | 1.778 | 2.167 | 1.778 | 0.000 |
| Qian (Wuhan) | 46 | China | 3.000 | 3.000 | 2.000 | 4.000 | 2.000 | 3.000 | 2.000 | 0.000 | 2.417 | 2.417 | 1.833 | 3.667 | 1.778 | 2.167 | 1.778 | 0.000 |
| Que | 47 | China | 3.000 | 3.000 | 2.000 | 4.000 | 1.000 | 2.720 | 2.000 | 3.000 | 3.000 | 3.000 | 2.000 | 4.000 | 0.692 | 2.462 | 2.000 | 3.000 |
| Saddik (General) | 48 | UAE | 3.000 | 1.800 | 1.877 | 3.077 | 0.985 | 1.800 | 1.400 | 3.800 | 2.797 | 1.620 | 1.544 | 2.532 | 0.810 | 1.481 | 1.152 | 3.658 |
| Saddik (Students) | 49 | UAE | 2.240 | 0.840 | 0.480 | 0.000 | 0.000 | 0.360 | 0.000 | 3.000 | 1.436 | 0.538 | 0.308 | 0.000 | 0.000 | 0.231 | 0.000 | 2.385 |
| Salman (Students) | 50 | Pakistan | 3.000 | 2.173 | 2.000 | 4.000 | 1.867 | 1.867 | 1.813 | 3.813 | 3.000 | 1.831 | 1.798 | 3.596 | 1.573 | 1.573 | 1.528 | 3.685 |
| Salman (Healthcare) | 51 | Pakistan | 3.000 | 2.280 | 2.000 | 4.000 | 2.000 | 2.000 | 1.960 | 4.000 | 3.000 | 2.203 | 2.000 | 4.000 | 1.844 | 1.844 | 1.781 | 3.953 |
| Sartorao Filho | 52 | Brazil | 3.000 | 3.000 | 2.000 | 4.000 | 2.000 | 1.962 | 2.000 | 4.000 | 3.000 | 3.000 | 2.000 | 3.850 | 2.000 | 1.625 | 2.000 | 4.000 |
| Shi | 53 | China | 3.000 | 3.000 | 2.000 | 4.000 | 2.000 | 3.000 | 2.000 | 1.280 | 3.000 | 3.000 | 2.000 | 4.000 | 2.000 | 2.951 | 2.000 | 0.780 |
| Sigdel | 54 | Nepal | 3.000 | 3.000 | 2.000 | 4.000 | 1.920 | 2.000 | 2.000 | 4.000 | 2.366 | 1.950 | 1.592 | 2.876 | 0.945 | 1.539 | 1.428 | 2.083 |
| Solomou | 55 | Cyprus | 3.000 | 2.000 | 2.000 | 3.429 | 1.619 | 1.619 | 1.619 | 4.000 | 2.400 | 1.429 | 1.771 | 2.057 | 0.971 | 0.971 | 0.971 | 3.086 |
| Stickley/Ueda | 56/57 | Japan | 3.000 | 1.000 | 1.000 | 0.000 | 0.000 | 0.632 | 1.000 | 2.842 | 3.000 | 1.000 | 1.000 | 0.000 | 0.000 | 0.387 | 1.000 | 2.516 |
| Stojanov (Healthcare/COVID) | 58 | Serbia | 3.000 | 3.000 | 2.000 | 4.000 | 2.000 | 3.000 | 2.000 | 4.000 | 3.000 | 3.000 | 2.000 | 4.000 | 2.000 | 3.000 | 2.000 | 4.000 |
| Stojanov (Healthcare/No-COVID) | 58 | Serbia | 3.000 | 3.000 | 2.000 | 4.000 | 2.000 | 3.000 | 2.000 | 4.000 | 3.000 | 3.000 | 2.000 | 4.000 | 2.000 | 3.000 | 2.000 | 4.000 |
| Sun | 59 | China | 3.000 | 2.778 | 2.000 | 4.000 | 1.222 | 2.833 | 1.917 | 2.444 | 3.000 | 2.840 | 2.000 | 4.000 | 1.440 | 2.880 | 1.940 | 2.160 |
| Tang W | 60 | China | 3.000 | 3.000 | 2.000 | 4.000 | 2.000 | 3.000 | 2.000 | 0.273 | 2.750 | 2.750 | 2.000 | 4.000 | 2.000 | 2.500 | 2.000 | 0.167 |
| Temsah | 61 | Saudi Arabia | 0.000 | 0.000 | 0.000 | 0.000 | 0.000 | 0.000 | 0.000 | 0.000 | 0.000 | 0.000 | 0.000 | 0.000 | 0.000 | 0.000 | 0.000 | 0.000 |
| Wang | 62 | China | 3.000 | 3.000 | 2.000 | 4.000 | 2.000 | 3.000 | 2.000 | 0.727 | 3.000 | 3.000 | 2.000 | 4.000 | 2.000 | 2.778 | 2.000 | 0.444 |
| Weilenmann | 63 | Switzerland | 3.000 | 2.591 | 2.000 | 3.591 | 0.000 | 0.864 | 0.864 | 3.000 | 1.917 | 1.583 | 2.000 | 2.583 | 0.000 | 0.528 | 0.528 | 1.917 |
| Xiao | 64 | China | 2.348 | 2.348 | 1.913 | 3.826 | 1.826 | 1.957 | 1.826 | 0.000 | 1.459 | 1.459 | 1.189 | 2.378 | 1.135 | 1.216 | 1.135 | 0.000 |
| Yamamoto | 65 | Japan | 3.000 | 1.000 | 1.000 | 0.000 | 0.000 | 1.000 | 1.000 | 3.000 | 3.000 | 1.000 | 1.000 | 0.000 | 0.000 | 1.000 | 1.000 | 3.000 |
| Zhang (Patient) | 66 | China | 3.000 | 3.000 | 2.000 | 4.000 | 2.000 | 3.000 | 2.000 | 0.000 | 2.273 | 2.273 | 1.758 | 3.515 | 1.697 | 2.000 | 1.697 | 0.000 |
| Zhang (Quarentine) | 66 | China | 3.000 | 3.000 | 2.000 | 4.000 | 2.000 | 3.000 | 2.000 | 0.000 | 2.273 | 2.273 | 1.758 | 3.515 | 1.697 | 2.000 | 1.697 | 0.000 |
| Zhang (General) | 66 | China | 3.000 | 3.000 | 2.000 | 4.000 | 2.000 | 3.000 | 2.000 | 0.000 | 2.273 | 2.273 | 1.758 | 3.515 | 1.697 | 2.000 | 1.697 | 0.000 |
| Zhao M | 67 | China | 2.182 | 2.182 | 1.818 | 3.636 | 1.727 | 1.773 | 1.727 | 0.000 | 1.333 | 1.333 | 1.111 | 2.222 | 1.056 | 1.083 | 1.056 | 0.000 |
| Zhao R | 68 | China | 3.000 | 3.000 | 2.000 | 4.000 | 2.000 | 2.500 | 2.000 | 0.000 | 1.853 | 1.853 | 1.471 | 2.941 | 1.412 | 1.588 | 1.412 | 0.000 |
| Zhou | 69 | China | 3.000 | 3.000 | 2.000 | 4.000 | 2.000 | 3.000 | 2.000 | 1.818 | 3.000 | 3.000 | 2.000 | 4.000 | 2.000 | 3.000 | 2.000 | 1.111 |
| Zhu Z | 70 | China | 2.824 | 2.824 | 2.000 | 4.000 | 2.000 | 2.176 | 2.000 | 0.000 | 1.548 | 1.548 | 1.290 | 2.581 | 1.226 | 1.258 | 1.226 | 0.000 |

**Table S4.** Previous prevalence of depression and anxiety based on the most recent published database of the Global Burden of Disease Study (GBD 2017 Disease and Injury Incidence and Prevalence Collaborators, 2018)

| Author | Ref | Country | Depression Cases*<br>(2017) | Anxiety Cases*<br>(2017) | Country<br>Population* (2017) | Depression<br>Prevalence (2017) | Anxiety Prevalence<br>(2017) |
| --- | --- | --- | --- | --- | --- | --- | --- |
| Ahmad | 11 | India | 30017 | 44873 | 1,338,659 | 2.24% | 3.35% |
| Ahn | 12 | Korea | 1222 | 2002 | 51,362 | 2.38% | 3.90% |
| Ahorsu (Female) | 13 | Iran | 3461 | 5754 | 80,674 | 4.29% | 7.13% |
| Ahorsu (Male) | 13 | Iran | 3461 | 5754 | 80,674 | 4.29% | 7.13% |
| Alyami | 14 | Saudi Arabia | 1257 | 1735 | 33,099 | 3.80% | 5.24% |
| Amerio | 15 | Italy | 1453 | 3379 | 60,537 | 2.40% | 5.58% |
| Bachilo | 16 | Russia | 3838 | 4398 | 144,497 | 2.66% | 3.04% |
| Bauer | 17 | Germany | 2043 | 5265 | 82,657 | 2.47% | 6.37% |
| Bauerle | 18 | Germany | 2043 | 5265 | 82,657 | 2.47% | 6.37% |
| Chang | 19 | China | 28672 | 44745 | 1,386,395 | 2.07% | 3.23% |
| Chen | 20 | China | 28672 | 44745 | 1,386,395 | 2.07% | 3.23% |
| Choi | 21 | China | 28672 | 44745 | 1,386,395 | 2.07% | 3.23% |
| Civantos | 22 | USA | 9611 | 20965 | 324,986 | 2.96% | 6.45% |
| Consolo | 23 | Italy | 1453 | 3379 | 60,537 | 2.40% | 5.58% |
| Fancourt | 24 | UK | 1809 | 2956 | 66,059 | 2.74% | 4.47% |
| Gao | 25 | China | 28672 | 44745 | 1,386,395 | 2.07% | 3.23% |
| Guo (Patient) | 26 | China | 28672 | 44745 | 1,386,395 | 2.07% | 3.23% |
| Hu | 27 | China | 28672 | 44745 | 1,386,395 | 2.07% | 3.23% |
| Islam | 28 | Bangladesh | 4294 | 6575 | 159,671 | 2.69% | 4.12% |
| Jia | 29 | UK | 1809 | 2956 | 66,059 | 2.74% | 4.47% |
| Johnson | 30 | Norway | 127 | 388 | 5,277 | 2.40% | 7.36% |
| Juanjuan | 31 | China | 28672 | 44745 | 1,386,395 | 2.07% | 3.23% |
| Kantor | 32 | USA | 9611 | 20965 | 324,986 | 2.96% | 6.45% |
| Kha. | 33 | India | 30017 | 44873 | 1,338,659 | 2.24% | 3.35% |
| Killgore | 34 | USA | 9611 | 20965 | 324,986 | 2.96% | 6.45% |
| Lai | 35 | China | 28672 | 44745 | 1,386,395 | 2.07% | 3.23% |
| Lin | 36 | China | 28672 | 44745 | 1,386,395 | 2.07% | 3.23% |
| Liu C | 37 | USA | 9611 | 20965 | 324,986 | 2.96% | 6.45% |
| Liu J | 38 | China | 28672 | 44745 | 1,386,395 | 2.07% | 3.23% |
| Mahedran | 39 | China | 28672 | 44745 | 1,386,395 | 2.07% | 3.23% |
| Mechili (Students) | 40 | Albania | 41 | 98 | 2,873 | 1.42% | 3.40% |
| Mechili (Family) | 40 | Albania | 41 | 98 | 2,873 | 1.42% | 3.40% |
| Munoz-.varro | 41 | Spain | 1824 | 2397 | 46,593 | 3.91% | 5.14% |
| Naser (General) | 42 | Jordan | 244 | 495 | 9,779 | 2.49% | 5.06% |
| Naser (Healthcare) | 42 | Jordan | 244 | 495 | 9,779 | 2.49% | 5.06% |
| Naser (Students) | 42 | Jordan | 244 | 495 | 9,779 | 2.49% | 5.06% |
| Nguyen | 43 | Vietnam | 1269 | 2006 | 94,597 | 1.34% | 2.12% |
| Olaseni | 44 | Nigeria | 4080 | 5341 | 190,873 | 2.14% | 2.80% |
| Pieh | 45 | Austria | 168 | 460 | 8,798 | 1.91% | 5.23% |
| Qian (Shangai) | 46 | China | 28672 | 44745 | 1,386,395 | 2.07% | 3.23% |
| Qian (Wuhan) | 46 | China | 28672 | 44745 | 1,386,395 | 2.07% | 3.23% |
| Que | 47 | China | 28672 | 44745 | 1,386,395 | 2.07% | 3.23% |
| Saddik (General) | 48 | UAE | 226 | 458 | 9,487 | 2.38% | 4.82% |
| Saddik (Students) | 49 | UAE | 226 | 458 | 9,487 | 2.38% | 4.82% |
| Salman (Students) | 50 | Pakistan | 3559 | 7345 | 207,897 | 1.71% | 3.53% |
| Salman (Healthcare) | 51 | Pakistan | 3559 | 7345 | 207,897 | 1.71% | 3.53% |
| Sartorao Filho | 52 | Brazil | 5031 | 13197 | 207,834 | 2.42% | 6.35% |
| Shi | 53 | China | 28672 | 44745 | 1,386,395 | 2.07% | 3.23% |
| Sigdel | 54 | Nepal | 948 | 1100 | 27,627 | 3.43% | 3.98% |
| Solomou | 55 | Cyprus | 24 | 66 | 1,180 | 2.05% | 5.61% |
| Stickley/Ueda | 56/57 | Japan | 2992 | 4305 | 126,786 | 2.36% | 3.40% |
| Stojanov (Healthcare/COVID) | 58 | Serbia | 182 | 318 | 7,021 | 2.59% | 4.53% |
| Stojanov (Healthcare/No-COVID) | 58 | Serbia | 182 | 318 | 7,021 | 2.59% | 4.53% |
| Sun | 59 | China | 28672 | 44745 | 1,386,395 | 2.07% | 3.23% |
| Tang W | 60 | China | 28672 | 44745 | 1,386,395 | 2.07% | 3.23% |
| Temsah | 61 | Saudi Arabia | 1257 | 1735 | 33,099 | 3.80% | 5.24% |
| Wang | 62 | China | 28672 | 44745 | 1,386,395 | 2.07% | 3.23% |
| Weilenmann | 63 | Switzerland | 204 | 446 | 8,452 | 2.41% | 5.28% |
| Xiao | 64 | China | 28672 | 44745 | 1,386,395 | 2.07% | 3.23% |
| Yamamoto | 65 | Japan | 2992 | 4305 | 126,786 | 2.36% | 3.40% |
| Zhang (Patient) | 66 | China | 28672 | 44745 | 1,386,395 | 2.07% | 3.23% |
| Zhang (Quarentine) | 66 | China | 28672 | 44745 | 1,386,395 | 2.07% | 3.23% |
| Zhang (General) | 66 | China | 28672 | 44745 | 1,386,395 | 2.07% | 3.23% |
| Zhao M | 67 | China | 28672 | 44745 | 1,386,395 | 2.07% | 3.23% |
| Zhao R | 68 | China | 28672 | 44745 | 1,386,395 | 2.07% | 3.23% |
| Zhou | 69 | China | 28672 | 44745 | 1,386,395 | 2.07% | 3.23% |
| Zhu Z | 70 | China | 28672 | 44745 | 1,386,395 | 2.07% | 3.23% |

*\*in thousands*

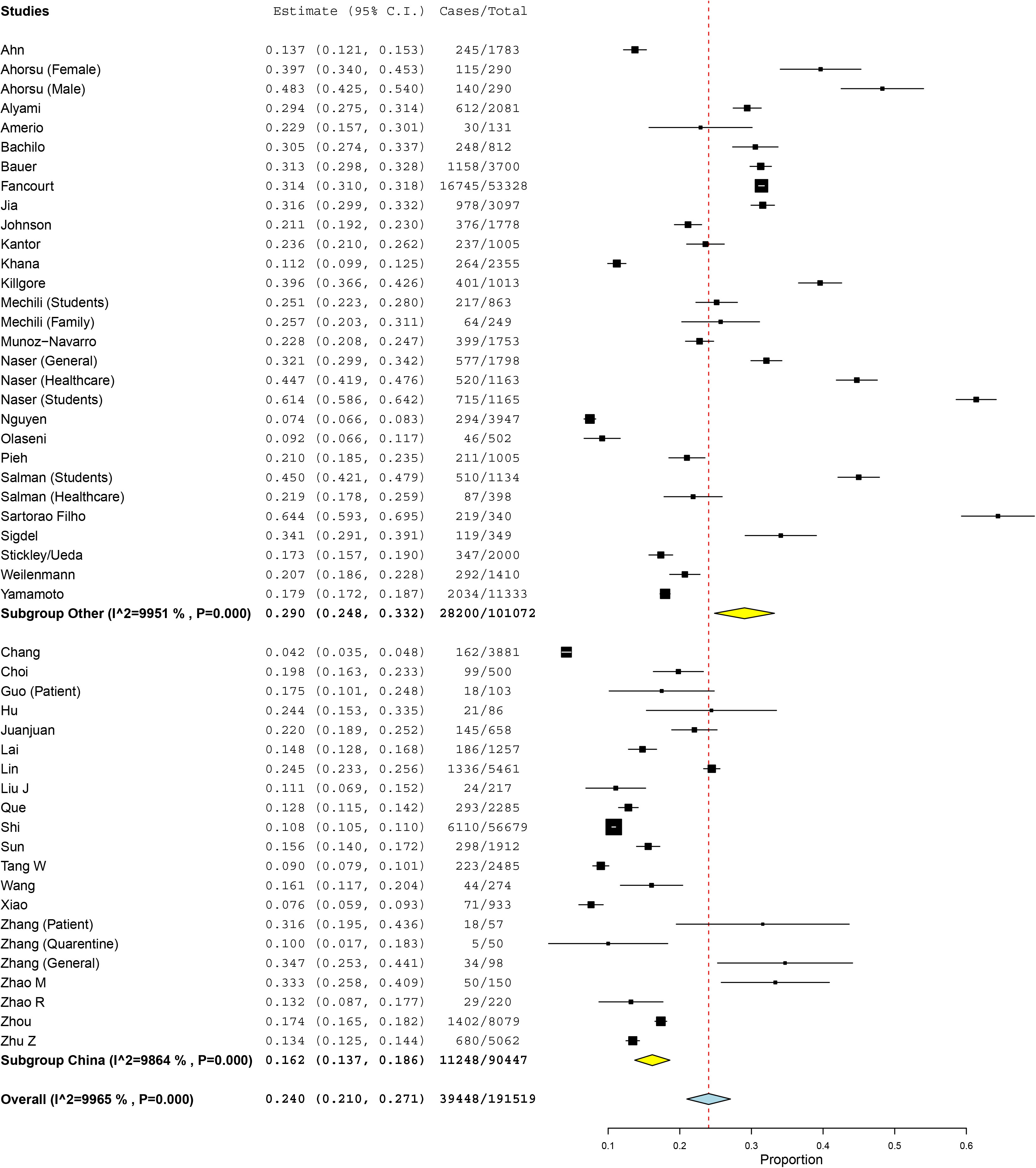
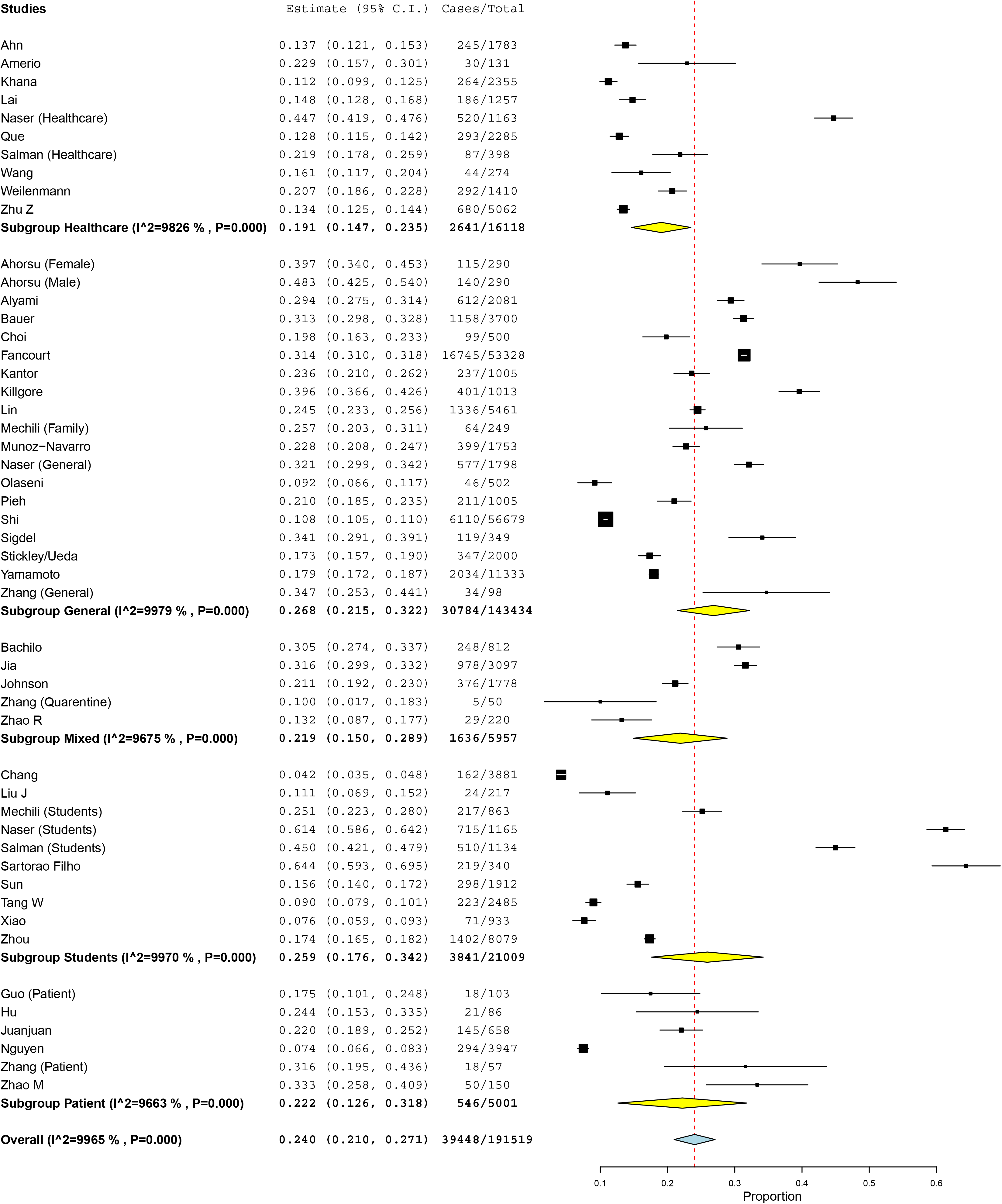
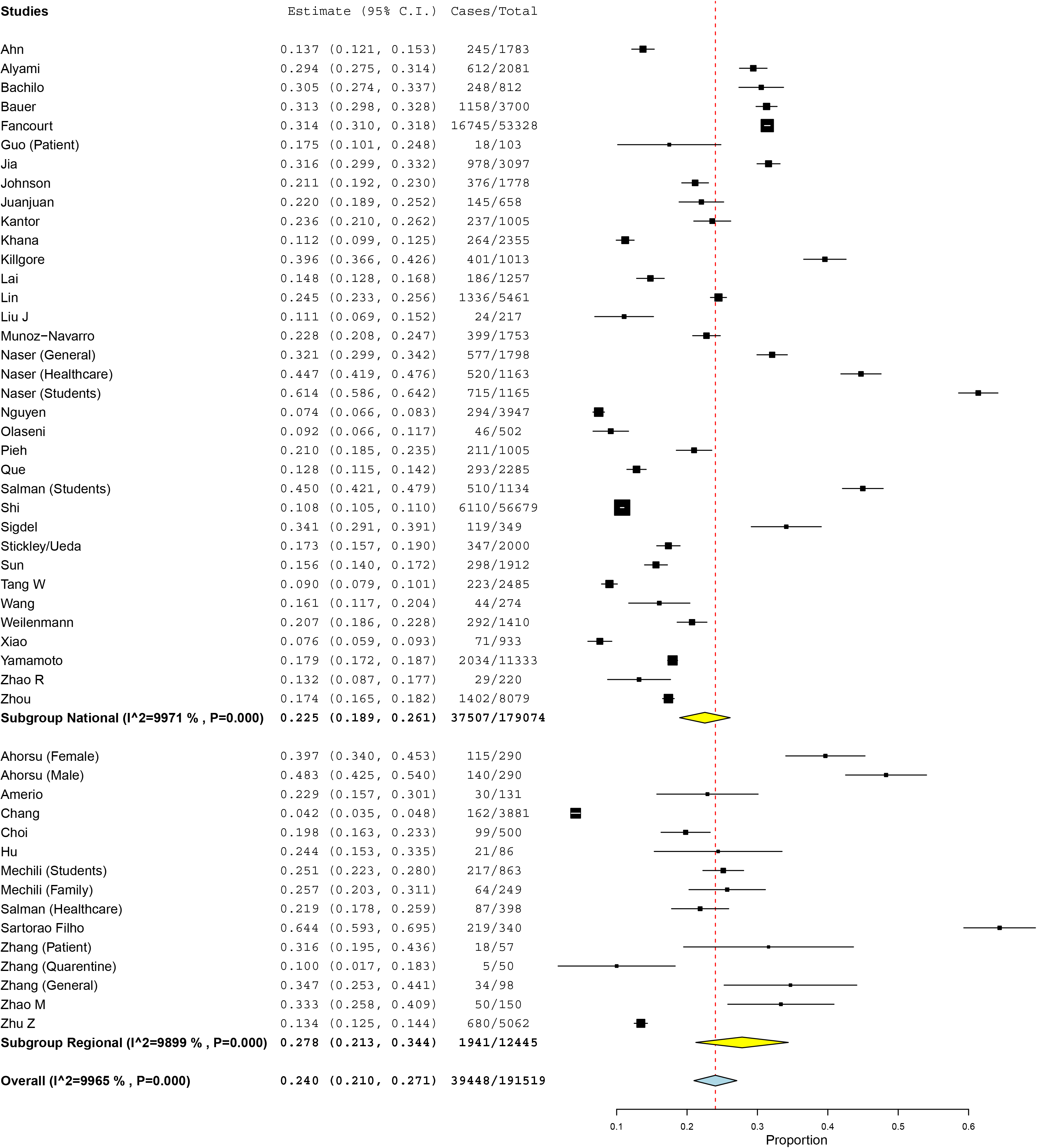
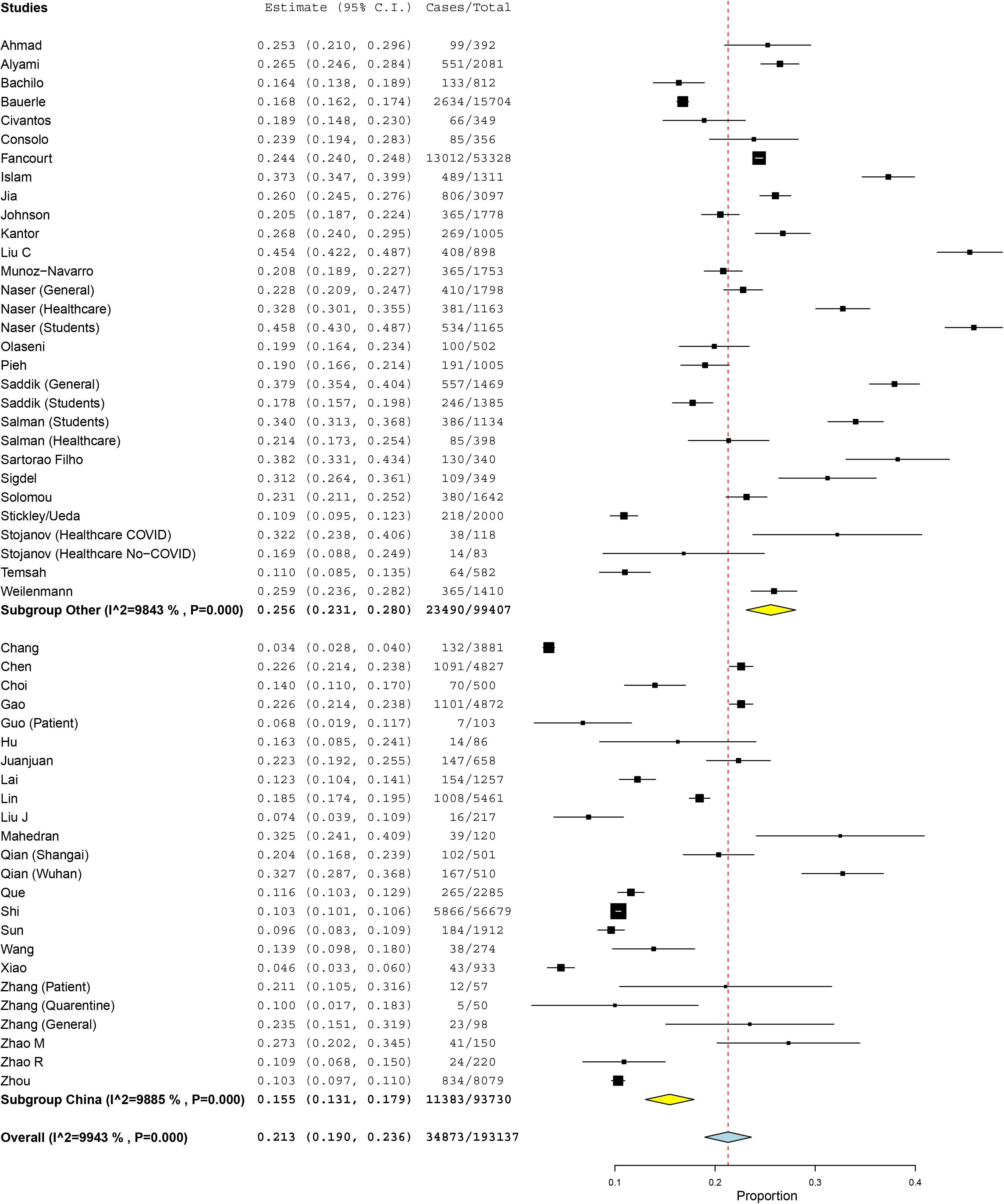
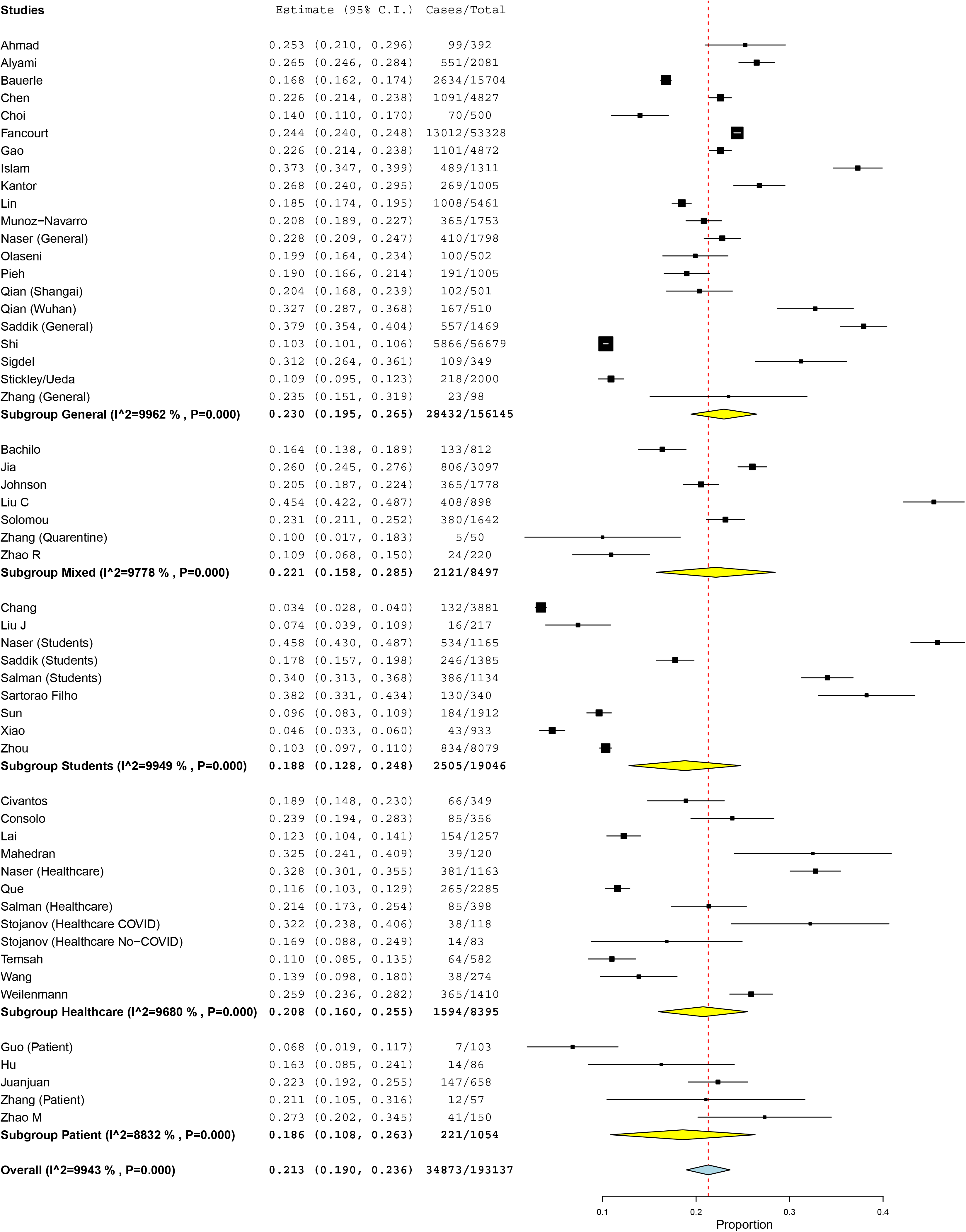
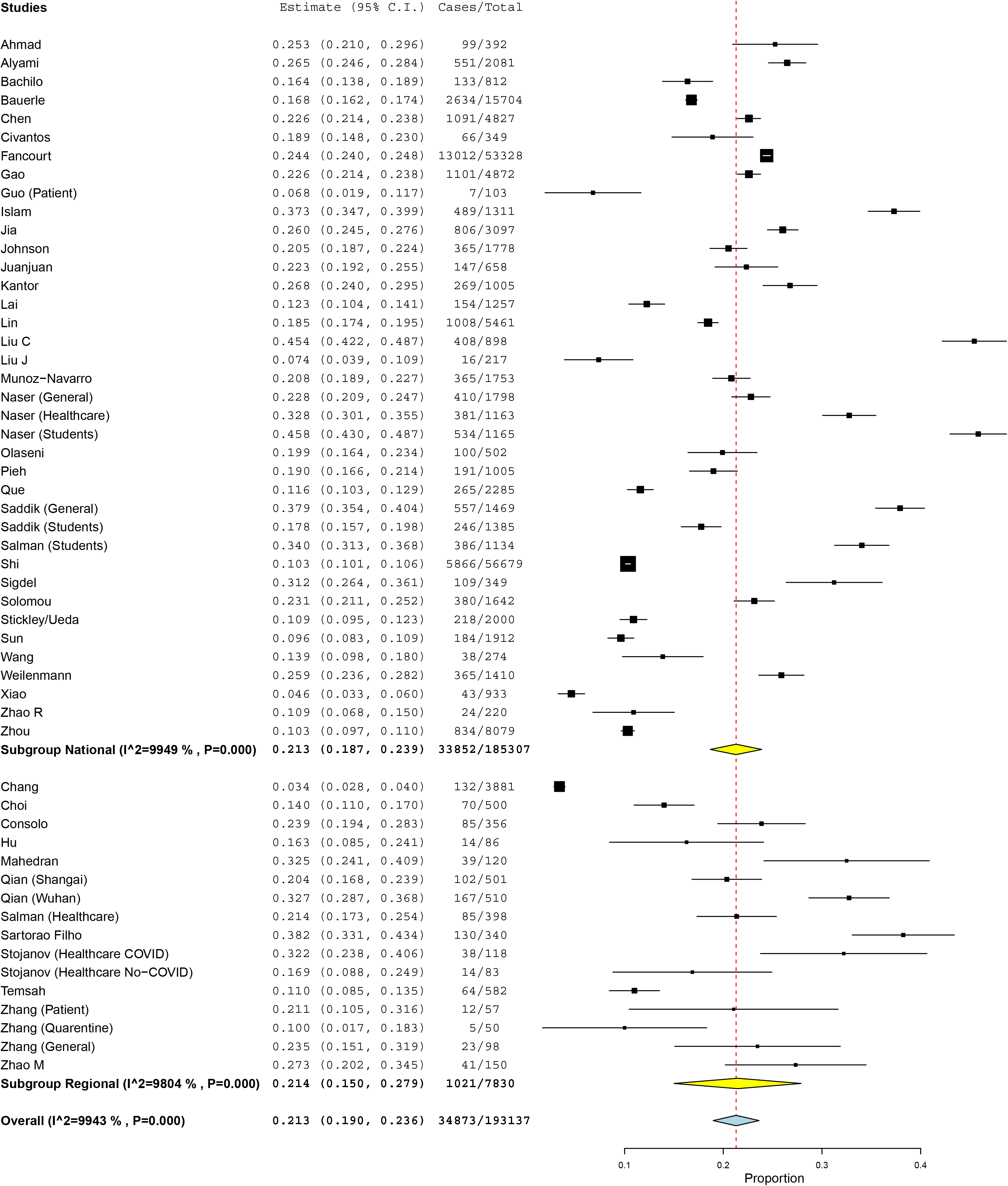

